# Cellular Diversity, Immune Crosstalk, and Genomic Alterations in Light Chain Amyloidosis

**DOI:** 10.1101/2025.06.06.25329039

**Authors:** Ankita Srivastava, Romika Kumari, Juho J. Miettinen, Minna H. Suvela, Klara Acs, Sini Luoma, Caroline A. Heckman

**Author notes:** Corresponding author: Dr. Caroline A. Heckman, Institute for Molecular Medicine Finland - FIMM, University of Helsinki, P.O. Box 20 (Tukholmankatu 8), 00014 University of Helsinki, Finland; Email address, Phone and Fax number. These authors contributed equally (Ankita Srivastava, Romika Kumari).

## Abstract

Plasma-cell heterogeneity in immunoglobulin light-chain amyloidosis (AL) drives disease variability and clinical outcomes and may be influenced by interactions with the cellular microenvironment, immune cell states, and light-chain (lambda/kappa) subtype. Concurrent multiple myeloma (MM) or precursor conditions (MGUS and sMM) introduce additional layers of cellular complexity. To characterize these processes across disease states, we performed single-cell RNA sequencing on bone marrow samples from 19 patients and identified 13 major cell types across 27 cell clusters, including six transcriptionally distinct plasma-cell subsets. Plasma-cell abundance negatively correlated with CD4+ and CD8+ T-cell levels, indicating reduced T-cell populations associated with high plasma-cell burden. Plasma cells exhibited 138 bidirectional ligand-receptor interactions across 35 significant pairs with 12 immune cell populations mediated through 15 pathways. The ligand/receptor interacting genes CXCR4 and PPIA from granulocyte-monocyte progenitors and monocyte subsets showed significant survival associations, underscoring the prognostic relevance of plasma cell-immune cell communication. T and NK cells exhibited variable exhaustion states, with higher exhaustion in CD4+ T-cells linked to poorer outcomes.

Differential expression analysis between kappa and lambda light chain subtypes plasma cells demonstrated increased inflammatory programs in kappa, while lambda plasma cells exhibited stronger unfolded protein response (UPR) and proliferative signatures. Across diagnostic groups, plasma cells formed a continuous transcriptomic spectrum; however, progression from AL-MGUS to AL-SMM to AL-MM was marked by loss of interferon-mediated inflammatory response alongside strong activation of MYC, p53, UPR, and apoptosis pathways, which were most prominent in AL-MM.

These findings elucidate key drivers of AL progression, stage/subtype vulnerabilities, revealed microenvironmental interactions, enabling improved patient stratification and targeted therapies.

**Key points:**

- Higher plasma-cell burden is linked to reduced CD4^+^/CD8^+^ T-cell proportions, with distinct exhaustion programs across CD4^+^, CD8^+^, and NK cells.
- Selective plasma cell-microenvironment communication networks shape disease biology, with specific ligand-receptor interactions linked to survival risk patterns.

## INTRODUCTION

Light-chain amyloidosis (AL) is a rare plasma cell (PC) disorder caused by extracellular deposition of misfolded immunoglobulin light chains in vital organs. The incidence of AL amyloidosis in the United States is estimated to be 16.7 per million persons per year with a prevalence of 69.0 per million adults^1^. Despite therapeutic advances, prognosis remains poor, with median survival ranging from 6 months to 3 years depending on organ involvement^2^.

AL amyloidosis arises from a clonal PC population that secretes unstable amyloidogenic lambda (λ) or kappa (κ) light chains, which misfold and assemble into fibrils that infiltrate organs such as the kidneys, heart, liver, gastrointestinal tract, and autonomic nervous system^3,4^. The light chain subtype is clinically relevant: λ AL is more prevalent and is associated with renal and neurologic involvement, whereas κ AL more often presents with hepatic disease^5^. Notably, patients with κ AL have significantly better progression free and overall survival despite similar treatment response rates, indicating that intrinsic biological differences between λ and κ clones may shape disease phenotype and outcome; however, the molecular basis for these disparities remains unclear^5^.

Cytogenetic studies have identified recurrent genomic abnormalities in AL amyloidosis. The most common events include t(11;14), hyperdiploidy with 1q gains, and monosomy 13/del(13q). Less frequent but prognostically adverse alterations such as del(17p)/*TP53* loss and t(4;14) mirror patterns observed in related plasma cell neoplasms, while t(14;16) is rare^6^. These findings underscore the genetic heterogeneity of AL PCs but do not fully explain organ tropism or light chain specific phenotypes.

T cells, especially CD8^+^ subsets, show activation and increased cytokine expression after daratumumab-based therapy in light chain amyloidosis, indicating enhanced anti-tumor responses^7^. NK cells express CD38 at lower levels than PCs, and anti-CD38 antibodies can modulate both PCs and immune cell populations including T and NK cells^8^. The precise mechanisms of PC interactions with T and NK cells in the bone marrow microenvironment are still poorly characterized, highlighting a key gap in knowledge^7,8^. Monocytes have been identified as a risk factor in AL amyloidosis, with predicted higher counts linked to increased disease risk and light chain clone selection.^9^ The findings indicate monocytes play roles in AL pathophysiology and progression, though mechanisms remain to be elucidated.

AL amyloidosis frequently co-occurs with other plasma cell dyscrasias such as multiple myeloma (MM; ∼10-20% of cases), monoclonal gammopathy of undetermined significance (MGUS), or smoldering multiple myeloma (SMM), reflecting shared clonal origins but distinct amyloidogenic potential. These associations complicate diagnosis and risk stratification^10^. The molecular mechanisms underlying the association between AL and other plasma cell dyscrasias remain unclear. Increased levels of bone marrow PCs in patients with AL have been previously associated with poor overall survival^11^. Patients with concurrent AL and MM are often excluded from clinical trials and tend to have poor prognosis and reduced overall survival^12,13^. Given that the approved standard of care is different for AL and other plasma cell dyscrasias, further investigation into the coexistence of these conditions is essential to enhance treatment strategies and to better understand the prognostic implications and characteristics of this association.

Single cell RNA sequencing (scRNA-seq) now enables high resolution characterization of inter and intra patient PC heterogeneity, bone marrow microenvironment interactions, and inference of copy number variation from transcriptomes. Although initial scRNA-seq studies of AL amyloidosis have highlighted limited PC transcriptional diversity and unique microenvironmental states^14^, several questions remain unresolved. In particular, λ *vs*. κ comparisons at single-cell resolution, AL-PC interactions with myeloid/lymphoid subsets via cell-cell communication, and differential expression in AL with co-occurring MM/MGUS/SMM are underexplored, limiting insights into progression from precursor states.

In this study, we applied scRNA-seq to bone marrow samples from patients with AL amyloidosis to systematically characterize plasma cell heterogeneity and light chain specific molecular features. We performed comparative analyses between λ and κ AL amyloidosis to identify transcriptional differences, plasma cell microenvironment interactions, and inferred copy number alterations. In addition, we compared AL PCs with co-occurring MM, SMM, and MGUS to delineate disease-specific transcriptional programs and genomic features.

## METHODS

### Sample collection

The Finnish Hematology Registry and Biobank (wwww.fhrb.fi) provided 20 samples of viably frozen bone marrow mononuclear cells (BM-MNCs) from 19 AL amyloidosis patients (n=19 at diagnosis; n=1 relapse sample). Diagnosis was confirmed via bone marrow (BM) biopsy with Congo red staining and amyloid typing (immunogold method or mass spectrometry) at the clinical diagnostic lab of Helsinki University Hospital (HUS Lab), and based on criteria established by the International Myeloma Working Group (IMWG)^15^ for co-occurring plasma cell dyscrasias. Light chain distribution among the cohort: 3 κ, 16 λ samples. Samples were subcategorized per BM PC aspirate/biopsy and IMWG CRAB criteria as AL-MGUS (n=3), AL-SMM (n=13), or AL-MM (n=3).

Staging was assessed using the Mayo Clinic 2004 system (troponin T >0.035 µg/L and NT proBNP >332 ng/L) and the 2012 revision, which additionally incorporated dFLC ≥180 mg/L. For κ, and λ restricted patients, dFLC was calculated as involved minus uninvolved light chain concentrations. Each marker above its cutoff (troponin T ≥25 ng/L, NT proBNP ≥1800 ng/L, dFLC ≥180 mg/L) contributed one point to assigning prognostic stage. Associated clinical data included cytogenetics via FISH. Samples were collected with informed consent per Helsinki University Hospital Ethics Committee approvals (permits 239/13/03/00/2010, 303/13/03/01/2011) and the Declaration of Helsinki.

### Single cell RNA-sequencing

Using fluorescence-activated cell sorting, the BM-MNCs were thawed and sorted based on cell viability using 7AAD and expression of the PC marker CD138 (APC labeled clone MI15 antibody from BD Biosciences, Santa Clara, CA). ScRNA-seq was performed using the Chromium Single Cell 3’ Gene Expression v3 reagent kit (10x Genomics, Pleasanton, CA). Gel beads in emulsion, cDNA amplification, and libraries were prepared following the manufacturer’s instructions. Between 4,000 and 9,000 live CD138-cells, along with the sorted CD138+ cells, were loaded per Chromium chip lane. Sequencing of the prepared libraries was performed using the Illumina NovaSeq 6000 system, which provided paired end reads of 28 bp and 89 bp.

### Data processing and analysis

The 10x Genomics Cell Ranger v6.0.2 pipelines were used for data processing and analysis. Specifically, ‘cellranger mkfastq’ was used to produce FASTQ raw sequence data files and ‘cellranger count’ was used to perform alignments, filtering and UMI (unique molecular identifier) counting. The alignments were performed against human genome GRCh38^16^. The output generated by the cellranger pipeline was used as input for the Seurat (version 5.1.0) tool^17^. Using Seurat, data quality control was performed, and the cells were selected for further analysis. Cells exhibiting low quality/dying cells and cell doublets or multiplets were filtered out using the filters nFeature_RNA > 200 and percent.mt cutoff was < triple median or 20%, whichever was the smaller value. After removing the unwanted cells, the data was normalized using the ‘LogNormalize’ method. Next, the top 2000 highly variable features were selected for downstream analysis. The batch effect in the data was corrected using the ‘rpca’ method. For linear dimension reduction, PCA was performed on the scaled data and the first 20 PCs were used in the ‘FindNeighbors’ function. A resolution of 0.8 and Louvain algorithm was used in ‘Findclusters’ function for clustering the cells. A non-linear dimensional reduction technique UMAP was used for visualization of the datasets. Differentially expressed genes were identified for all the clusters using ‘FindAllMarkers’ function with min.pct=0.25 and logfc.threshold=0.25. To assign cell type identity to the clusters, we used Azimuth^18^, and mononuclear cell markers were obtained from sctype database^19^. To recognize the heterogeneity and similarities among the various cell populations, we investigated the enriched biological processes within these cell types using Escape v2.1.0^20^. Cell-cell communication was analyzed using CellChat v2.2.0 with the full CellChatDB (Secreted Signaling, Cell-Cell Contact, ECM-Receptor, Non-Protein Signaling). Probabilities were computed from ligand-receptor expression for network construction and visualization. This quantified intercellular signaling pathways from single-cell transcriptomic data^21^.

The immune exhaustion landscape was assessed using established markers of chronic dysfunction: inhibitory receptors (*PD-1, LAG-3, HAVCR2/TIM-3, TIGIT, CTLA-4), NK-specific NKG2A/CD94 (KLRC1/KLRD1*), killer receptors (*CD160, CD244, CD96*), and transcriptional programs (*TOX, EOMES, IKZF2, ZNF683/Hobit*) identified in our cohort across CD8⁺, CD4⁺, and NK cell populations^22–25^. Cell-type-specific exhaustion scores were then calculated using AddModuleScore (Seurat) for CD8⁺ T cells (*PDCD1, LAG3, HAVCR2, TIGIT, CTLA4, TOX, EOMES, IKZF2, CD244, CD160, ZNF683*), CD4⁺ T cells (*PDCD1, LAG3, TIGIT, CTLA4, TOX, ENTPD1, IKZF2, HAVCR2, CD244*), and NK cells (*KLRC1, KLRD1, TIGIT, LAG3, CD96, ZNF683, HAVCR2, TOX, CD244, CD160*), followed by median stratification.

Differential expression analysis of PCs was performed to compare light chain subtypes (κ *vs*. λ samples) and disease subcategories (AL-MGUS, AL-SMM and AL-MM samples), focused on autosomal genes using Seurat’s single-cell and pseudobulk methods. Genes with consistent significance (p_val.bulk < 0.05 and p_val.sc < 0.05) across both methods were defined as differentially expressed (DE), classified as highly upregulated if avg_log2FC.sc > 0.5 and avg_log2FC.bulk > 0.5, or highly downregulated if avg_log2FC.sc < −0.5 and avg_log2FC.bulk < −0.5. Comparative pathway analysis between AL subtype and disease category was done using ‘Single cell pathway analysis’ (SCPA)^26^.

To identify somatic large-scale chromosomal copy number alterations including gains and deletions of entire chromosome or large segments of the chromosomes, we used InferCNV^27^. For inferCNV, we used the following cell types: natural killer (NK) cells, CD8⁺ T cells, CD4⁺ T cells, CD14^+^ and CD16^+^ monocytes, late erythrocytes, B cells, granulocyte monocyte progenitor (GMP), hematopoietic stem cells (HSC), conventional (cDC2) and plasmacytoid dendritic cells (pDC) and proliferating cells (non-tumor) and plasma cell as malignant cell type. The parameters we used included cutoff=0.1, cluster_by_groups=T, denoise=T and HMM=T. Further, we utilized the output from the inferCNV analysis, particularly the expression data matrix found in the expr.data slot of the inferCNV object (infercnv_obj@expr.data), to combine the inferCNV results from 20 samples. The expression data matrix contains the processed and normalized gene expression values for all cells in each sample. Each gene in the matrix was mapped with its specific cytoband based on its location in the genome. For each sample, we followed these steps; a) we grouped genes according to their cytobands; b) we calculated the average expression value for all genes within each cytoband.

## RESULTS

### Clinical, cytogenetic and cardiac biomarker landscape of the study cohort

To gain a deeper understanding of the cellular diversity and molecular mechanisms associated with AL we analyzed clinical data from 19 patients contributing 20 samples (diagnosis: n=19; relapse: n=1). The cohort included 11 male and 8 female participants, with a median age of 64 years and median survival time of 40.4 months. The cohort included both λ and κ light chain amyloidosis (λ: n=16; κ: n=3) and exhibited various cytogenetic aberrations, with 1q gain (5/19), t(11;14)(5/19) and trisomy 11(4/19) being the most frequent abnormalities (**Figure 1B, Supplemental Figure 1(A) and Table 1**), in line with previous reports^28^.

**Figure 1.**
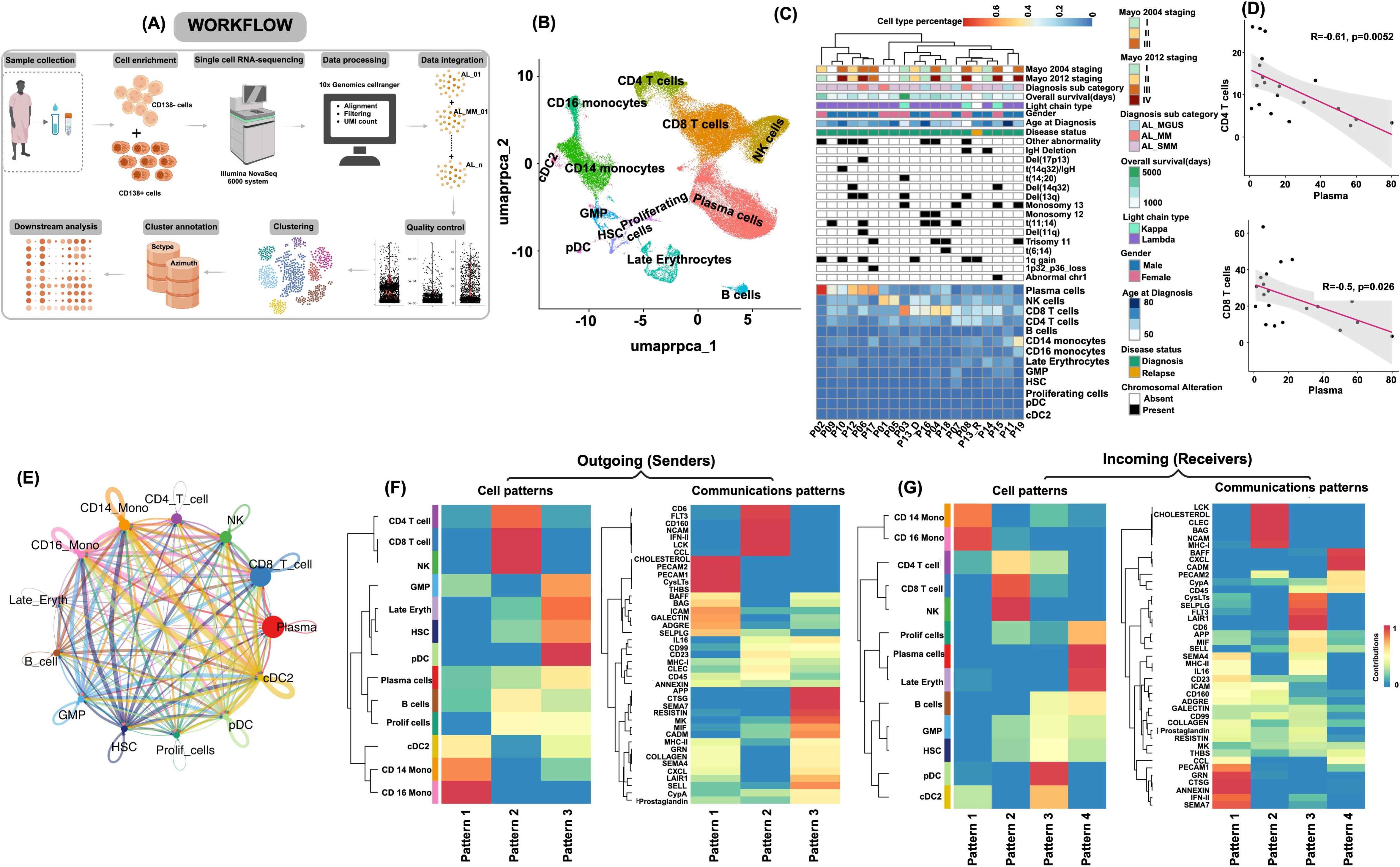
Overview of Study Design and Single-Cell Landscape of AL Amyloidosis Revealing Plasma Cell–Driven T-Cell Depletion and Distinct Survival-Signaling and Cell–Cell Communication Patterns. (A) A flowchart illustrating the steps of the study design, which includes sample collection, single-cell RNA sequencing, and subsequent analysis. The figure was created on BioRender.com. (B) RPCA-based integration of 20 samples (19 patients + 1 relapse) identifies 13 distinct immune and hematopoietic cell types across 27 clusters in light chain amyloidosis. UMAP projection of aggregated cells (n = 101, 227) from 19 patients (+ 1 relapse)samples, with each dot representing the transcriptome of an individual cell and a color code indicating distinct clusters. (C) A heatmap showing the clinical and cellular overview of the cohort (n = 20), including diagnosis, overall survival (days), light-chain type, demographic information, cytogenetic aberrations, and Mayo 2004/2012 staging, together with the corresponding cell-type proportions. The bottom heatmap displays the percentage of different predicted cell-type populations in the 20 samples. (D) Inverse correlation between plasma cell abundance and T cell populations. (E) Aggregated cell–cell communication network across all cell types in light chain amyloidosis, visualized as a CellChat circle plot. Node size represents the relative abundance of each cell group, and edge thickness reflects the number of inferred interactions. (F) Identification of communication patterns among cell types using CellChat. Outgoing communication patterns resolved by NMF reveal three distinct themes. Heatmaps show the contribution of each cell type and signaling pathway to the outgoing patterns. (G) Incoming communication patterns identified by NMF, showing four major patterns. Heatmaps illustrate the contribution of cell types and signaling pathways to incoming patterns.

**Table 1.** Individual patient characteristics in the AL amyloidosis study cohort.

| Patient id | Disease status | Age at the time of diagnosis | Overall survival days | Os status | Gender | Light chain type | FISH Findings | BM plasma cell % in aspirate (morphology) | BM plasma cell % in biopsy | Serum M component g/l | crabroti | Final category | Serum Kappa FLC mg/l | Serum Lambda FLC mg/l | dFLC | proBNP ng/l | TnT ng/l | Mayo Clinic 2012 revision | Mayo 2004 Staging |
| --- | --- | --- | --- | --- | --- | --- | --- | --- | --- | --- | --- | --- | --- | --- | --- | --- | --- | --- | --- |
| P01 | Diagnosis | 66-70 | 196 | 1 | Female | lambda | None | 8 | 30 | 0 | 00A0 | AL_MM | 13 | 8190 | 8177 |  |  |  |  |
| P02 | Diagnosis | 61-65 | 2945 | 0 | Female | lambda | 1q_gain\|other_abnormality\|11 | 10 | 15 | 11,8 |  | AL_SMM | 6,3 | 61,7 | 55,4 | 1469 | 23 | I | II |
| P03 | Diagnosis | 51-55 | 5381 | 0 | Female | kappa | t(14_20)\|del(13q)/-13 | 8 | 7 | 22,2 |  | AL_SMM | 72,1 | 7,2 | 64,9 | 37 | 6 | I | I |
| P04 | Diagnosis | 61-65 | 2580 | 0 | Female | lambda | t(11_14)\|trisomy_11\|other_abnormality\|monosomy_12 | 15 | 20 | 0 |  | AL_SMM | 11,3 | 487 | 475,7 | 5209 | 42 | IV | III |
| P05 | Diagnosis | 71-75 | 1229 | 0 | Female | lambda | None | <5 | 12 | 0,0 |  | AL_SMM | 32,6 | 426,0 | 393,4 |  | 88,0 |  |  |
| P06 | Diagnosis | 61-65 | 449 | 1 | Male | lambda | del(13q)\|del(17p13)\|t(11_14)\|1q_gain\|other_abnormality\|del(11q) | 49 | 45 | 3,5 | 00AB | AL_MM | 22,9 | 1140,0 | 1117,1 | 7356 | 128,0 | IV | III |
| P07 | Diagnosis | 51-55 | 789 | 0 | Male | lambda | t(11_14)\|monosomy_13 | 6 |  | 0,4 |  | AL_MGUS | 20,1 | 35,0 | 14,9 | 95 |  |  |  |
| P08 | Diagnosis | 45-50 | 764 | 0 | Female | kappa | del(13q)\|1q_gain\|other_abnormality\|IgH_deletion | 25 | 80 | 0,5 | 000B | AL_MM | 2890,0 | 6,5 | 2883,5 | 29620 | 132,0 | IV | III |
| P09 | Diagnosis | 66-70 | 253 | 1 | Male | lambda | t(11_14) | 10 | 13 | 4,2 |  | AL_SMM | 80,6 | 284,0 | 203,4 | 9188 |  |  |  |
| P10 | Diagnosis | 71-75 | 691 | 0 | Male | lambda | 1q_gain\|other_abnormality\|IgH_(14q32)_translocation | 8 | 13 | 0 |  | AL_SMM | 10,1 | 277,0 | 266,9 | 11074 | 95,0 | IV | III |
| P11 | Diagnosis | 81-85 | 787 | 1 | Male | lambda | NA | 5 |  | 2,2 |  | AL_MGUS | 33,3 | 63,9 | 30,6 | 2548 |  |  |  |
| P12 | Diagnosis | 61-65 | 1924 | 0 | Male | lambda | del(13q)\|other_abnormality\|del(14q32) | 16 |  | 0,5 |  | AL_SMM | 15,8 | 273 | 257,2 | 715 | 10 | II | II |
| P13_D | Diagnosis | 66-70 | 1805 | 0 | Male | lambda | 1q_gain | 50 | 25 | 3,7 |  | AL_SMM | 20,6 | 791 | 770,4 | 838 | 16 | II | II |
| P13_R | Relapse | 66-70 |  |  | Male | lambda | 1q_gain |  |  |  |  | AL_SMM |  |  |  | 463 | 17 |  | II |
| P14 | Diagnosis | 71-75 | 1838 | 0 | Male | lambda | IgH_deletion | 4 | 10 | 4,5 |  | AL_SMM | 31,8 | 32,8 | 1 | 169 | 6 | I | I |
| P15 | Diagnosis | 61-65 | 39 | 1 | Female | kappa | monosomy_13\|del(14q32)\|structurally_abnormal_chromosome_1 | 6 | 15 | 1,1 |  | AL_SMM | 562 | 15,9 | 546,1 | 9300 | 237 | IV | III |
| P16 | Diagnosis | 76-80 | 1664 | 0 | Male | lambda | t(11_14)\|other_abnormality\|monosomy_12 | 7 | 7 | 1 |  | AL_MGUS | 17 | 41,5 | 24,5 | 37 | 8 | I | I |
| P17 | Diagnosis | 66-70 | 1604 | 0 | Male | lambda | 1p32/p36_loss\|trisomy_11 | 20 | 30 | 10,9 |  | AL_SMM | 11,4 | 119 | 107,6 | 8558 | 269 | III | III |
| P18 | Diagnosis | 56-60 | 1632 | 0 | Female | lambda | t(6_14)\|trisomy_11 | 25 | 20 | 3 |  | AL_SMM | 9,3 | 164 | 154,7 | 191 | 6 | I | I |
| P19 | Diagnosis | 61-65 | 153 | 1 | Male | lambda | trisomy_11\|monosomy_13 | 10 | 15 | 0 |  | AL_SMM | 29,1 | 287 | 257,9 | 9773 | 163 | IV | III |

\*Mayo 2004 Score: 1 point each for proBNP ≥ 332 ng/L and TnT ≥ 35 ng/L; total score translates to stage (0 = I, 1 = II, 2 = III).
Mayo 2012 Score: 1 point for each of dFLC ≥ 180 mg/L, proBNP ≥ 1800 ng/L, and TnT ≥ 25 ng/L; total score translates to stage (0 = I, 1 = II, 2 = III, 3 = IV).

Further, the hematological parameters which were reported for these patients included serum free light chain median κ concentration (serum κ FLC) of 20.1 mg/l (range: 6,3-2890 mg/l) and median λ concentration (serum λ FLC) of 91.45 mg/l (range: 6,5-8190 mg/l). The corresponding immunoglobulin light chain values (S-IgLcK-V and S-IgLcL-V) were consistent with these findings. The median difference between the involved and uninvolved light chains (dFLC) was 257,2 mg/l (range: 1-8177 mg/l). Serum M-component was detected in ∼73.68% of patients with a median concentration of 1.65 g/l (range: 0-22,2 g/l). The median BM PC infiltration on aspirate morphology was 10% (range: 4-50%) and 15% (range: 7-80%) on biopsy sections. Cardiac biomarkers were elevated in most patients with median proPNP of 2008,5 ng/l (range: 37-29620 ng/l)a nd median Troponin T (TnT) of 16.5 ng/l (range: 6-269 ng/l). The above-mentioned biomarkers were used for staging: Mayo 2004^29^ [NTproBNP ≥ 332 ng/l, TnT ≥ 35 ng/l]:- stage I (n=4), II (n=3), III (n=7), missing(n=5), with the relapse sample remaining at stage II; Mayo 2012 revised^30^ [dFLC ≥ 180 mg/l, NTproBNP ≥ 1800 ng/l, TnT ≥ 25-ng/l]: stage I (n=5), II (n=2), III (n=1), IV (n=6), missing (n=6). Overall, the observed parameters showed significant clonal PC burden and frequent cardiac involvement in the patients (**Table 1**).

### The immune microenvironment in AL amyloidosis

We analyzed 20 scRNA-seq samples (19 diagnostic, 1 relapse), yielding 148,224 cells. Each cell had a median of 36,202 reads and 1,237 genes detected prior to quality filtering. After filtering, 101,227 high quality cells remained for downstream analyses. From this filtered dataset, we identified 27 transcriptionally defined clusters corresponding to 13 major cell populations (**Figure 1B-C, Table S2**).

These clusters were annotated according to the expression of known cell type biomarkers, as shown in **Supplemental Figure 1B** (see the methods section for additional details). The identified cell types included PCs, NK cells, CD8⁺ T cells, CD4⁺ T cells, CD14^+^ and CD16^+^ monocytes, late erythrocytes, B cells, GMP, HSC, cDC2 and pDC and proliferating cells (**Figure 1B-C**). The cell types that exhibited more than one cluster included PCs (n=6), CD8⁺ T cells (n=4), CD4⁺ T cells (n=3), late erythrocytes (n=3), CD14^+^ monocytes (n=2) and NK cells (n=2). This clustering of the same cell-type in more than one cluster can be attributed to factors like cell states and biological heterogeneity^31^.

Further, PCs were the most abundant cell type among the obtained clusters constituting 29.8% of the total cells, as expected given that CD138^+^ cell enrichment was performed prior to sequencing, followed by CD8⁺ T cells (27.8%), NK cells (11.1%), CD14^+^ monocyte cells (9.7%). Hierarchical clustering of cell type proportions revealed two subclusters: one enriched for PCs and another with higher NK and CD8⁺ T cell levels. Most samples in the PC-rich cluster were Mayo 2004 stage II or higher, with a similar pattern in Mayo 2012, except for sample P02, which was classified as stage I only in the Mayo 2012 system **(Figure 1C)**.

To further investigate the tumor microenvironment in AL, we examined the correlation between the proportion of different predicted cell types. We found that the proportions of CD4⁺ (R = –0.61, p = 0.0052) and CD8⁺ T cells (R = –0.50, p = 0.026) were negatively correlated with the proportions of PCs (**Figure 1D and Supplemental Figure 1C**). These findings suggest that increased PC burden is associated with reduced T cell presence, indicating potential immune suppression or altered immune dynamics in AL amyloidosis. A strong correlation between other cell types was also observed, as shown in **Supplementary Figure 1C**. Lymphocytes (including CD8⁺ T cells, CD4⁺ T cells, NK cells, B cells and PCs) and myeloid cells (including CD16^+^ monocytes, CD14^+^ monocytes, late erythroid, GMP, HSC, pDC cDC2 and proliferating cells) were clearly distinguishable based on the enriched biological states. One of these biological processes was the ‘KRAS_SIGNALING_DN’ gene set, which includes genes that are downregulated by ‘KRAS activation’, and this gene set was significantly enriched in the lymphocytes (**Supplemental Figure 1D and E**).

### Global communication mapping identifies signaling patterns and cell type–specific pathways in AL amyloidosis

CellChat analysis revealed dynamic intercellular communication among all 13 cell types in light chain amyloidosis. The most interactive cells were cDC2 (544 interactions), CD14^+^ monocytes (498), CD16^+^ monocytes (454), and pDCs (358), indicating their central role in shaping the immune microenvironment, while PCs showed selective signaling interactions; **Figure 1E**).

Directional signaling highlighted cDC2, CD14^+^ monocytes, HSCs, and GMPs as top senders, and CD14^+^/CD16^+^ monocytes, CD8^+^ T cells, and cDC2 as top receivers; PCs had 93 outgoing and 55 incoming (148 total) interactions. Ten candidate interactions were identified in the count matrix but excluded later in the analysis due to lack of statistical significance (p ≥ 0.05) **(Supplementary Table 3)**.

Global pattern analysis identified 41 active signaling pathways out of 290 curated in CellChat, spanning immune regulatory pathways (MHC I, MHC II, MIF, GALECTIN, IFN II, IL16), adhesion-related (ICAM, PECAM1/2, SELPLG, SELL), extracellular matrix remodeling (COLLAGEN, THBS, APP), chemokine signaling (CCL, CXCL), and others including MK, BAFF, CypA, Prostaglandin, ADGRE, and ANNEXIN. The optimal number of communication patterns was determined based on outgoing and incoming signaling profiles across all annotated cell types in our samples. It revealed three distinct outgoing (sender) patterns and four incoming (receiver) patterns, effectively grouping cell types by shared signaling behaviors for clearer network visualization (**Figure 1F&G**). PCs contributed to all three outgoing patterns, with strongest involvement in Pattern 3 which is enriched for APP, CTSG, SEMA7A, RETN, MK, MIF, CADM, SELL, LAIR1, and IL16 (while other pathways overlapped more with Patterns 1/2). For incoming profiles, PCs aligned primarily with Pattern 4 (shared with proliferating and late erythroid cells), enriched for BAFF, CXCL, CADM, PECAM2, CypA, and CD45 pathways tied to immune regulation, adhesion, and survival. Notably, strong BAFF levels have also been linked to autoimmune hemolytic anemia^32^. **Supplementary Figure 1F** shows cell type-specific signaling strength. This highlights the selective role of PCs in receiving immune activation and tissue interaction signals (**Figure 1F-G, Supplementary Figure 1F**).

### Plasma cell signaling hubs and prognostic interactions in AL amyloidosis

Of 41 pathways observed in the AL amyloidosis immune landscape, 15 were significantly active in PCs (all p < 0.05), with 138 of 148 total detected interactions reaching statistical significance. PC-centered communication analysis revealed extensive crosstalk across the microenvironment, with PCs interacting most prominently with CD8^+^ T cells (n = 14), CD16^+^ monocytes (n = 12), CD14^+^ monocytes (n = 11), cDC2 (n = 10), NK cells (n = 8), and GMPs (n = 8), indicating broad engagement with both cytotoxic lymphocytes and myeloid populations **(Figure 2A)**. These 138 interactions spanned 88 outgoing and 50 incoming signals across 35 unique ligand–receptor pairs.

**Figure 2.**
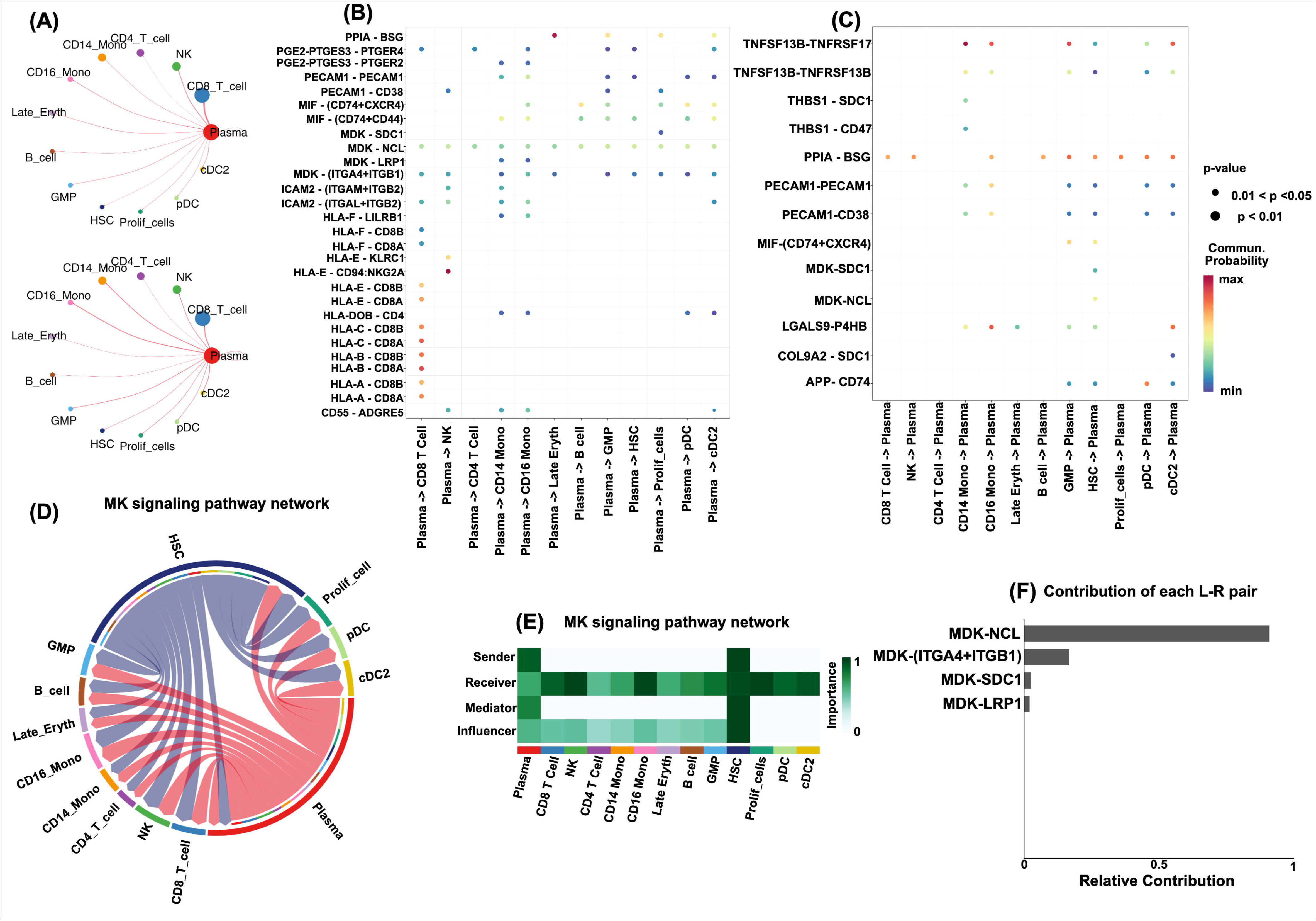
Plasma-centered cell–cell communication analysis highlighting the MK signaling axis. **(A)** Circle plot showing the number of interactions and interaction weights involving plasma cells across all major cell types. **(B)** Bubble plot showing significant ligand–receptor interactions sent from plasma cells signaling to other cell populations. **(C)** Bubble plot showing significant ligand–receptor interactions received by plasma cells. **(D)** Chord diagram illustrating the MK signaling network, in which plasma cells and HSCs emerge as the dominant sending populations. **(E)** Heatmap of MK signaling centrality scores demonstrating that CD8 T cells, NK cells, CD16^+^ monocytes, proliferating cells, cDC2, and pDC are the major receiving populations. **(F)** Bar plot shows the relative contribution of ligand–receptor pairs within the MK pathway, identifying MK–NCL as the most prominent interaction.

Outgoing signaling was mediated through 10 pathways including MK, MHC-I, MIF, Prostaglandin, ICAM, PECAM1, CypA, MHC-II, ADGRE, and PECAM2. The MK pathway dominated (n = 25) via MDK–NCL and MDK–ITGA4/ITGB1(VLA4), followed by MIF–(CD74+CD44), MIF–(CD74+CXCR4), and PGE2–PTGER4 to monocytes, HLA-A/B/C/E:CD8A to CD8^+^ T cells, and HLA-E:CD94-NKG2A to NK cells, suggesting pathways through which PCs reprogram myeloid cells for tumor support and suppress NK cytotoxicity (Figure 2B, D–F). Incoming signals originated predominantly from HSC (n = 10), cDC2 (n = 8), GMP (n = 8), and CD14^+^/CD16^+^ monocytes (n = 13), mediated through BAFF, CypA, GALECTIN, PECAM1, PECAM2, APP, MIF, MK, THBS, and COLLAGEN. BAFF signaling dominated (n = 12) via TNFSF13B–TNFRSF17 and TNFSF13B–TNFRSF13B from GMP, HSC, and monocytes, with PPIA–BSG (n = 9) and LGALS9–P4HB (n = 6) also prominent, indicating mechanisms by which PCs receive and integrate external cues from the immune microenvironment **(Figure 2C)**. CypA, MIF, MK, PECAM1, and PECAM2 functioned bidirectionally **(Supplementary Figure 1F)**.

We further examined signaling directionality and key ligand–receptor pairs across MIF, BAFF, CypA, and PECAM pathways, revealing multiple significant PC-involving interactions that collectively reflect the complex interplay of immune signaling, adhesion, and matrix remodeling in AL amyloidosis **(Supplementary Figure 2A–D, Tables 6–7)**.

To assess clinical relevance, we extracted all significant ligand–receptor pairs specific to PC interactions with each cell type including GMP, CD14^+^/CD16^+^ monocytes, NK cells, CD8^+^ T cells, and CD4^+^ T cells (low-abundance populations cDC2 and pDC excluded) and evaluated the corresponding ligand and receptor genes in Cox proportional hazards and Kaplan–Meier survival analyses. Among all cell-type-specific L-R genes examined, CXCR4 in GMPs and PPIA in CD16^+^ monocytes emerged as significant prognostic markers. Higher CXCR4 expression in GMPs was strongly associated with poorer survival (Cox HR 9.8, 95% CI 1.1–87.1, p = 0.04; log-rank p = 0.009), while higher PPIA in CD16^+^ monocytes showed a consistent prognostic trend (Cox HR 2.88, 95% CI 1.03–8.03, p = 0.043; log-rank p = 0.091), though the latter did not reach significance on Kaplan–Meier analysis, warranting validation in larger cohorts **(Supplementary Figure 2E–F)**.

### Diversity and oncogenic-prognostic signatures of plasma-cells in AL amyloidosis

A total of 6 PC clusters (PC_01-PC_06) were identified from 30,191 plasma cells (n=20 patients, 101,227 total cells) and annotated using canonical markers (**Supplemental Figure 1B; Figure 3A**). Furthermore, these clusters revealed inter- and intra-patient heterogeneity with uniform distribution across samples (**Figure 3B**) Analysis of gene expression within these clusters revealed up-regulation of various gene sets, including immunoglobulin genes **(Figure 3C)**. Further, we examined the association of the top 10 highly expressed genes in each cluster with overall survival. Among all genes tested, *IGHA1* showed a significant association with survival in the Cox proportional hazards model (HR 1.98, 95% CI 1.04–3.76, p=0.037). Although Kaplan–Meier plot based on median *IGHA1* expression stratification showed a trend toward differential overall survival, it did not reach statistical significance (log-rank p = 0.089). **Supplementary Figure 3A-B**). Additionally, pathway enrichment analysis revealed PC_01 and PC_05 as closely related, PC_02 and PC_04 as transcriptionally similar, and PC_03 and PC_06 as the most distinct clusters (**Figure 3D**).

**Figure 3.**
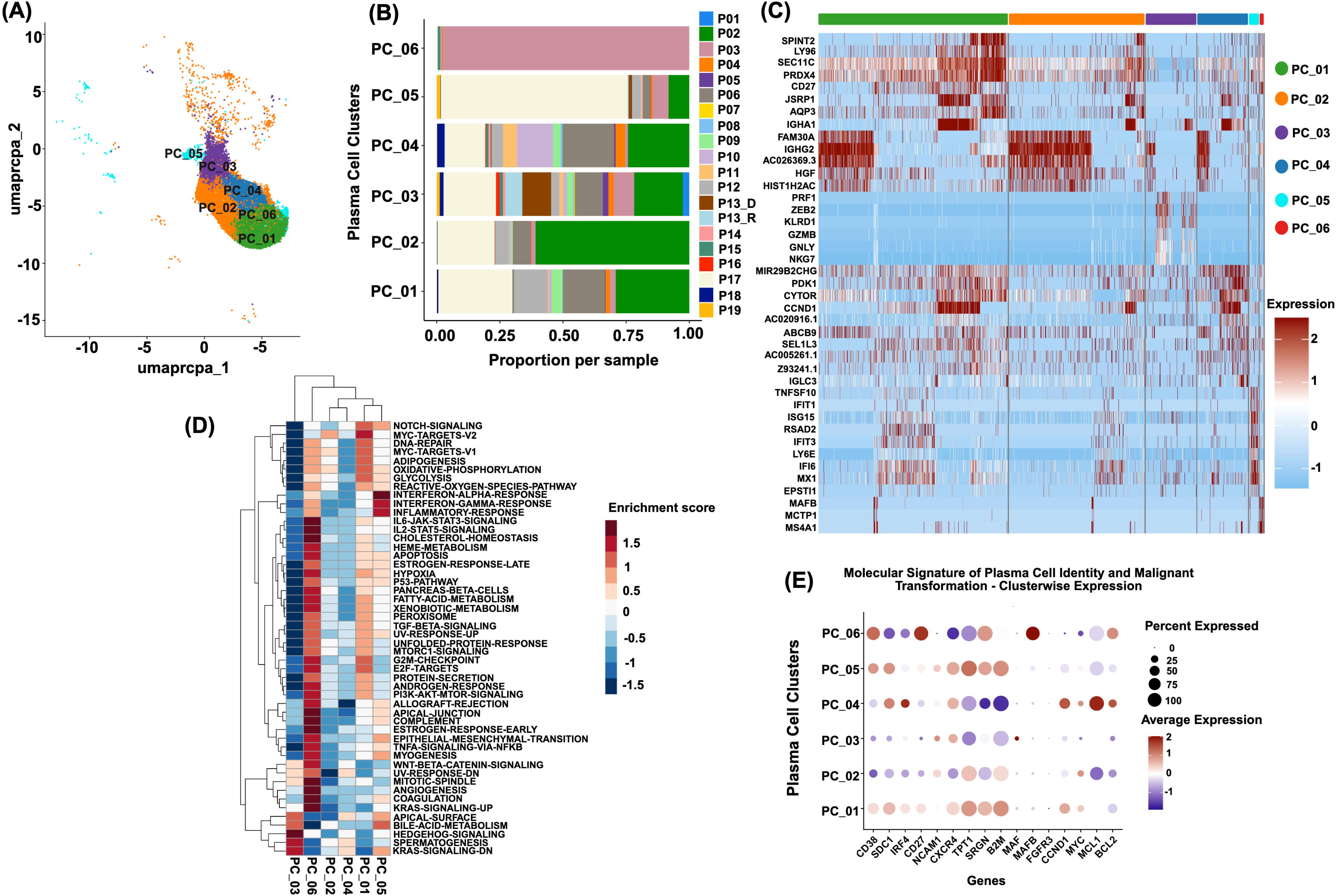
Plasma Cell Clusters Reveal Transcriptional Heterogeneity, Shared Pathways, and Oncogenic–Prognostic Signatures in AL Amyloidosis. **(A)** UMAP visualization of six plasma cell clusters, with each dot representing an individual plasma cell (n= 30,191 cells). **(B)** The composition of plasma cells within each plasma cell cluster across the 20 patient samples in the cohort. **(C)** A heatmap displaying the top 10 differentially expressed genes for each plasma cell cluster (n = 6), where each column corresponds to a plasma cell and each row represents a gene; the top bar indicates the different plasma cell clusters, each marked in distinct colors. **(D)** Pathway enrichment analysis of plasma cell clusters, highlighting the similarities and differences among different plasma cell clusters. **(E)** Dot plot visualization of plasma cell identity and malignant transformation associated gene signatures demonstrates differential expression patterns across plasma cell clusters, indicating distinct functional states within the malignant plasma cell compartment.

Genes defining malignant transformation, including PC identity/surface markers & microenvironment interaction (*CD38, SDC1, CD27, NCAM1, B2M, CXCR4, SRGN*), core transcriptional/cell-cycle oncogenic drivers (*IRF4, MYC, CCND1, MAF*, *MAFB, FGFR3*), and survival/apoptosis-resistance programs (*MCL1, BCL2, TPT1*), were curated from established MM transcriptomic signatures with cluster-specific expression examined in the our PC cluster^33,34^. PC_04 exhibited elevated *CCND1, BCL2,* and *MCL1*, suggesting an apoptosis-resistant malignant clone. PC_01, PC_02, and PC_05 showed higher *B2M* and *TPT1*, suggesting tumor burden/stress-adaptation; PC_01, PC_05, and PC_06 displayed elevated *SRGN*, suggesting stroma-remodelling. PC_06 expressed high *MAFB* and *BCL2*, marking a MAF-translocated, survival-adapted subtype. These patterns reveal distinct malignant plasma cell subclusters relevant to AL amyloidosis progression (**Figure 3E**).

### Cellular and transcriptomic changes from diagnosis to relapse

The samples from patient P13 were collected at both diagnosis and relapse stages. A comparison of the cell-type proportions between these stages revealed an increase in PCs (1.8%), indicating a higher tumor burden at relapse. Additionally, we observed an increase in the proportions of CD4^+^ T cells (13.5%), CD16^+^ monocytes (4.19%), late erythrocytes (3.5%), GMP (0.4%), HSC (0.26%), pDC (0.1%), and cDC2 (0.3%). Conversely, the proportions of NK cells (9.57%), CD8^+^ T cells (9.39%), CD14^+^ monocytes (0.55%), B cells (3.9%), and proliferating cells (0.6%) were found to be lower at relapse **(Supplementary Figure 3C)**. However, we did not observe much heterogeneity within the transcriptome profiles of the PC populations between diagnosis and relapse, indicating tumor clonal evolution from diagnosis to relapse **(Supplementary Figure 3D)**. Differential gene expression analysis of PCs from the paired samples identified 21 genes with altered regulation. The diagnosis sample showed upregulation of the following genes: *IFI44L, S100A9, XAF1, MX1, GIMAP7, IFI44, IFI6, GIMAP4, CHST2, HERC6* and *IL1B*. Meanwhile, the genes *IGLC2, IGHD, BTG3, PER1, ENC1, U2AF1, GRASP,* and *IGLV3-1* were downregulated **(Supplementary Figure 3E)**. Given that the paired sample size in this study was limited to just one pair, these findings should be interpreted with caution and should be further validated in a larger cohort.

### T cell and NK cell subsets display distinct exhaustion signatures with prognostic implications

Single-cell analysis identified four CD8⁺ T, three CD4⁺ T, and two NK cell clusters with distinct gene expression patterns and cell frequencies **(Figure 4A)**. To further characterize immune cell functional states and identify exhausted subpopulations, we examined exhaustion marker expression within these clusters, revealing variable levels across CD8⁺, CD4⁺, and NK clusters (**Methods; Figure 4B**). CD4⁺ T cells in clusters 2 and 3 exhibited moderate but consistent *ENTPD1, IKZF2,* and *CTLA4* expression, reflecting partial exhaustion. NK cells across both clusters showed high exhaustion signatures (*HAVCR2, TIGIT, TOX, EOMES, KLRC1, KLRD1, CD244, CD160, CD96*). CD8⁺ T cells displayed ubiquitous *LAG3* expression across all four clusters, with *TIGIT* upregulated in clusters 3-4, *KLRD1* in 1, 2, and 4, CD96 in 1-3, *ZNF683/Hobit* in 1-2, and rare *PDCD1/TOX* cells throughout (**Figure 4B**). Enrichment of canonical T-cell-exhaustion genes in these subpopulations suggests an exhausted phenotype, in line with prior studies in hematologic malignancies^35^.

**Figure 4.**
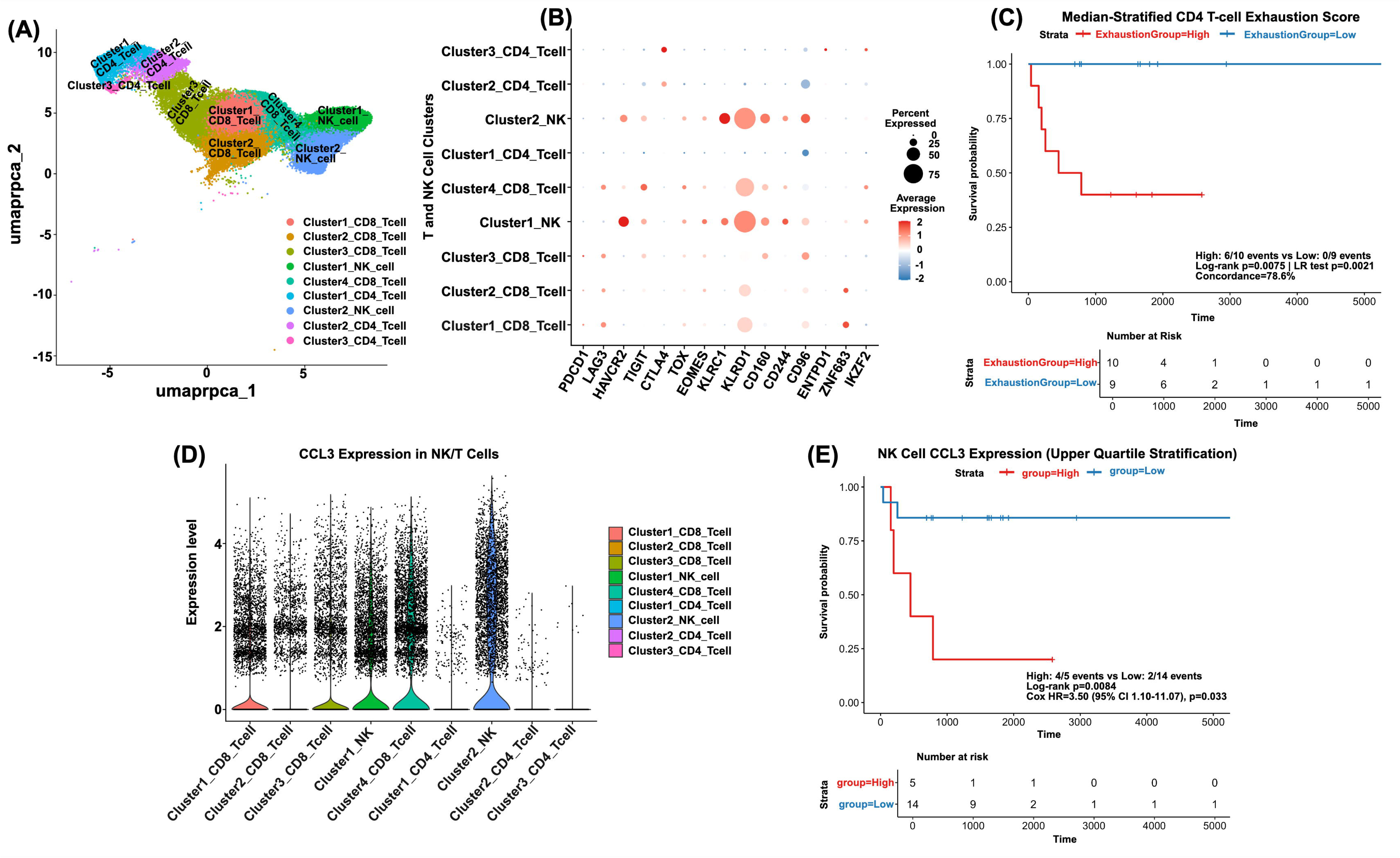
Identifying Trends of Lymphocyte Dysfunction and Their Potential Impact on Survival in AL. **(A)** UMAP representation of lymphocyte subclusters, including CD4⁺ T-cell, CD8⁺ T-cell, NK cell populations. **(B)** Dot plot showing the expression of exhaustion- and dysfunction-related genes across lymphocyte subclusters. **(C)** Kaplan–Meier survival analysis stratified by median CD4⁺ T-cell exhaustion score, demonstrating poorer survival in patients with high exhaustion states. **(D)** Violin plot showing the expression levels of CCL3 across the NK-cell cluster and other lymphocyte subsets. **(E)** Kaplan–Meier curve based on upper-quartile stratification of NK-cell CCL3 expression, showing significantly reduced survival associated with high CCL3-expressing NK populations.

Despite rare PDCD1⁺ CD8⁺ T cells (primarily P03, P13_D, P19), median-stratified high expression predicted significantly worse survival (KM log-rank p=0.034; Supplementary Figure 4A-B). Distinct CD8⁺, CD4⁺, and NK cell subsets showed elevated exhaustion scores despite low baseline expression in most cells (**Supplementary Figure 4D**). Cell-type-specific exhaustion scores (Methods) with median stratification showed prognostic value for overall survival using CD4⁺ T-cell exhaustion (High: n=9, 6/6 events; Low: n=10, 0/10 events; log-rank p=0.0075, LR test p=0.0021, concordance=78.6%) (**Figure 4C**).

The top 15 markers per CD8⁺, CD4⁺, and NK cell cluster were screened for overall survival correlation. CCL3, highly expressed in NK cells, showed prognostic significance: upper quartile stratification had 4/5 events in the high group (log-rank p=0.008; Cox HR=1.69, p=0.033), while median was non-significant (p=0.15) (**Figure 4D & E**). Given the small cohort size (n=19), statistical power was limited. Larger studies are needed to validate CD4⁺ T-cell exhaustion and NK cell *CCL3* expression as prognostic biomarkers.

### Distinct transcriptomic signatures define κ and λ AL amyloidosis plasma cell subtypes

To investigate molecular differences between κ (n=3) and λ (n=16) AL amyloidosis subtypes at the plasma cell level, differential gene expression analysis was performed. UMAP projection revealed spatial separation with partial intermingling, indicating transcriptional divergence (**Figure 5A**). Cluster distribution showed strong light chain bias: PC_01-PC_05 were 90-98% λ, while PC_06 was ∼99% κ (**Figure 5B**). Differential expression identified 107 genes (FDR < 0.05, |log2FC| > 0.5), with 29 upregulated (such as, *IGKC, KLF1, RHPN1, TCN2, FN1, HBG2, CD200R1, CCL3*) and 78 were downregulated in κ in comparison to λ subtype (*CCND1, IGLC2/IGLC7, JCHAIN, NCAM1, DKK1, SYT1, PAX5, ANGPT2, BANK1*) samples (**Figure 5C**). Single-cell pathway analysis (SCPA) confirmed distinct pathway activity: κ cells showed strong enrichment in allograft rejection (q=8.17, FC=9.05), interferon gamma response (q=7.84, FC=7.81), TNFα/NFκB signaling (q=7.84, FC=14.56), apoptosis (q=7.67, FC=5.64), and epithelial–mesenchymal transition (q=7.67, FC=2.02), while they exhibited suppression of MYC targets V1 (q=6.4, FC=-16.8), unfolded protein response (UPR) (q=6.7, FC=-5.6), and oxidative phosphorylation (q=5.3, FC=-18.9) (**Figure 5D-E**). Conversely, λ cells showed enrichment of MYC target and metabolic pathways with reduced inflammatory and apoptotic signaling, indicating a more quiescent, metabolically active state.

**Figure 5.**
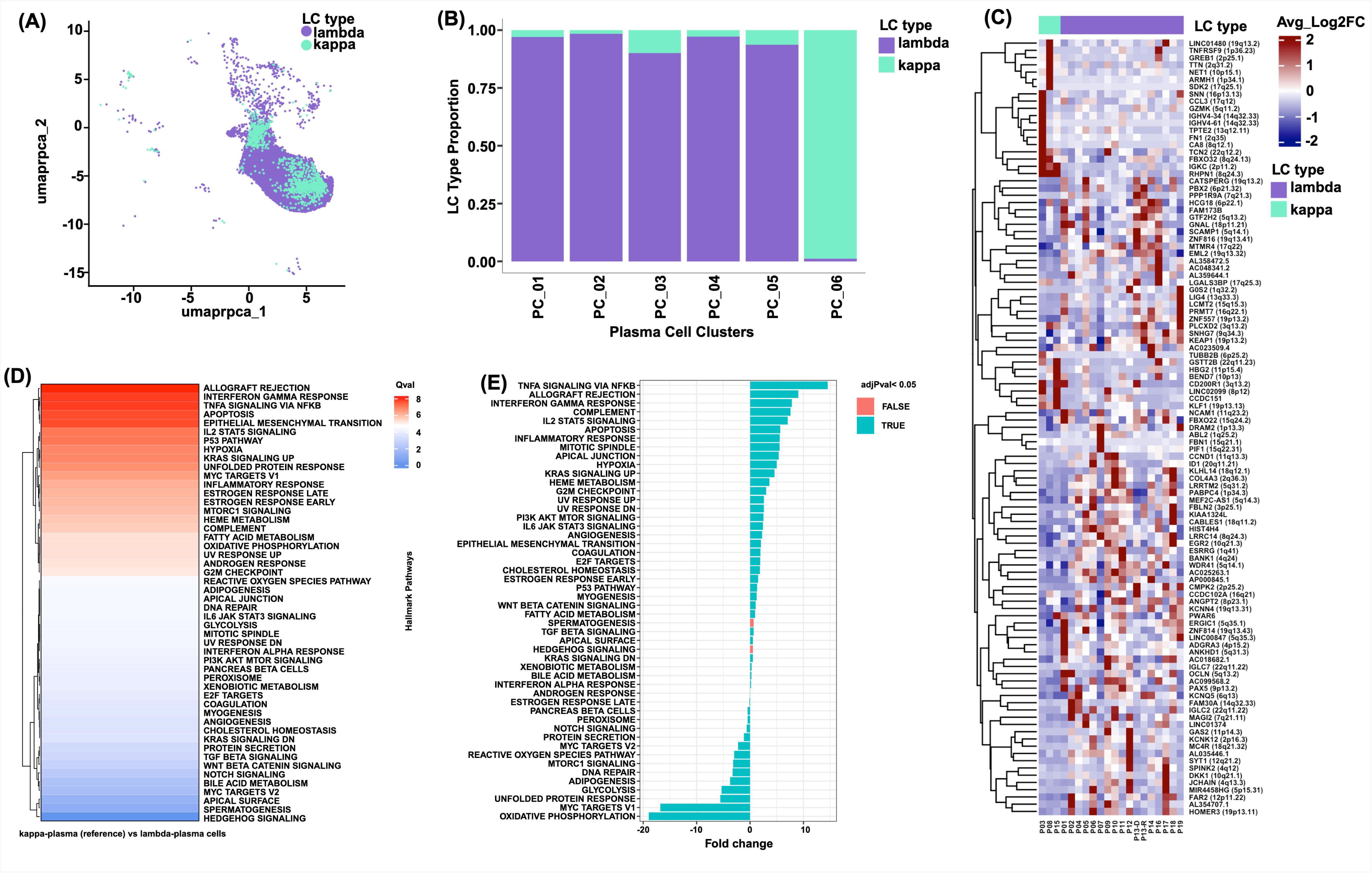
Transcriptomic and Pathway Divergence Between Kappa and Lambda Plasma Cells. **(A)** UMAP projection of plasma cells annotated by light-chain type (lambda vs. kappa). **(B)** Proportion of lambda and kappa plasma cells across each plasma-cell cluster. **(C)** Heatmap illustrating the differentially regulated genes between lambda and kappa plasma cells. **(D)** Relative pathway enrichment analysis of plasma cell populations in kappa vs lambda samples using Single Cell Pathway Analysis (SCPA), where q-values shown in the heatmap serve as the primary metric; higher q-values indicate greater divergence in pathway activity between conditions. **(E)** Diverging bar chart displaying fold-change (FC) enrichment scores computed between the two plasma-cell subtypes (kappa vs lambda).

### The MYC, p53, and apoptosis pathways underlies the progression from AL MGUS/SMM to AL_MM

Based on IMWG criteria, we classified samples as AL_MM (n=3), AL_SMM (n=13), and AL_MGUS (n=3). Plasma cells from all groups intermingled in UMAP space, occupying overlapping regions with broadly similar transcriptional profiles across diagnostic categories (**Supplementary Figure 5A**). AL_SMM cells dominated all six plasma cell clusters (73-100%), with minor AL_MM (up to 21% in PC_04) and AL_MGUS (0-6%) contributions, largely reflecting sample composition (**Supplementary Figure 5B**).

DE analysis using both bulk and single-cell approaches (p < 0.05, |log₂FC| > 0.5) identified 107 differentially regulated genes between AL_MM and AL_MGUS, with 68 upregulated and 39 downregulated in AL_MM in comparison to AL_MGUS (**Supplementary Figure 5C**). SCPA revealed top pathways by q-value including TNFα signaling via NF-κB (q=7.85, FC=-1.22), apoptosis (q=7.59, FC=3.61), and interferon gamma response (q=7.24, FC=-9.96), alongside pathways with extreme fold-changes such as MYC targets V1 (q=6.54, FC=32.42), oxidative phosphorylation (q=5.74, FC=27.99), and p53 (q=7.15, FC=9.72) in AL_MM compared to AL_MGUS (**Supplementary Figure 5D-E**). AL_MM *vs*. AL_SMM showed 159 upregulated and 309 downregulated genes in AL_MM (Supplementary Figure 5F; **Supplementary Table10**). SCPA (AL_MM vs AL_SMM) showed top q-value pathways TNFα signaling via NF-κB (q=8.59, FC=8.39), apoptosis (q=8.42, FC=4.55), and p53 (q=8.17, FC=3.55), alongside extreme fold-changes in oxidative phosphorylation (q=5.49, FC=-10.56), allograft rejection (q=6.58, FC=-7.04), and interferon gamma response (q=8.00, FC=-6.49) (**Supplementary Figure 5G-H**). In contrast, differential gene expression between AL_MGUS and AL_SMM plasma cells identified 8 upregulated and 462 downregulated genes in AL_MGUS (Supplementary Figure 5I). SCPA (AL_MGUS *vs*. AL_SMM) showed top q-value pathways TNFα signaling via NF-κB (q=7.67, FC=7.88), allograft rejection (q=7.06, FC=-7.18), and p53 (q=6.97, FC=-7.38), alongside extreme fold-changes in oxidative phosphorylation (q=6.54, FC=-39.54), MYC targets V1 (q=6.45, FC=-33.49), and mTORC1 signaling (q=6.10, FC=-7.08) (**Supplementary Table 11, Supplementary Figure 5J-K**). These findings reveal hyperactivated stress/pro-survival signaling with suppressed metabolic and inflammatory programs in AL_MM relative to the precursor states.

### InferCNV uncovers distinct genomic alterations within AL

Somatic large-scale chromosomal copy number alterations (CNAs) were identified using inferCNV^27^, considering PCs as malignant and other cell types as non-tumor references. PCs exhibited widespread CNAs across the cohort, with patient P12 showing deletions on chromosomes 13p, 14, and 16. Recurrent gain of chromosome 1q was detected in PC subclusters from patients P2, P6, and P10^36^ (**Supplementary Figure 6**).

Unsupervised clustering of all samples (n = 20) defined distinct CNV groups, reflecting inter-patient heterogeneity. Cluster 1 showed higher overall CNV burden, while cluster 2 was enriched for 14q gain (P3, P9, P11, P18) and chromosome 22 amplification (P4, P9, P10, P11, P18). Integration with clinical annotations (disease status, cytogenetics, stage, survival) revealed no clear associations (**Figure 6A**).

**Figure 6:**
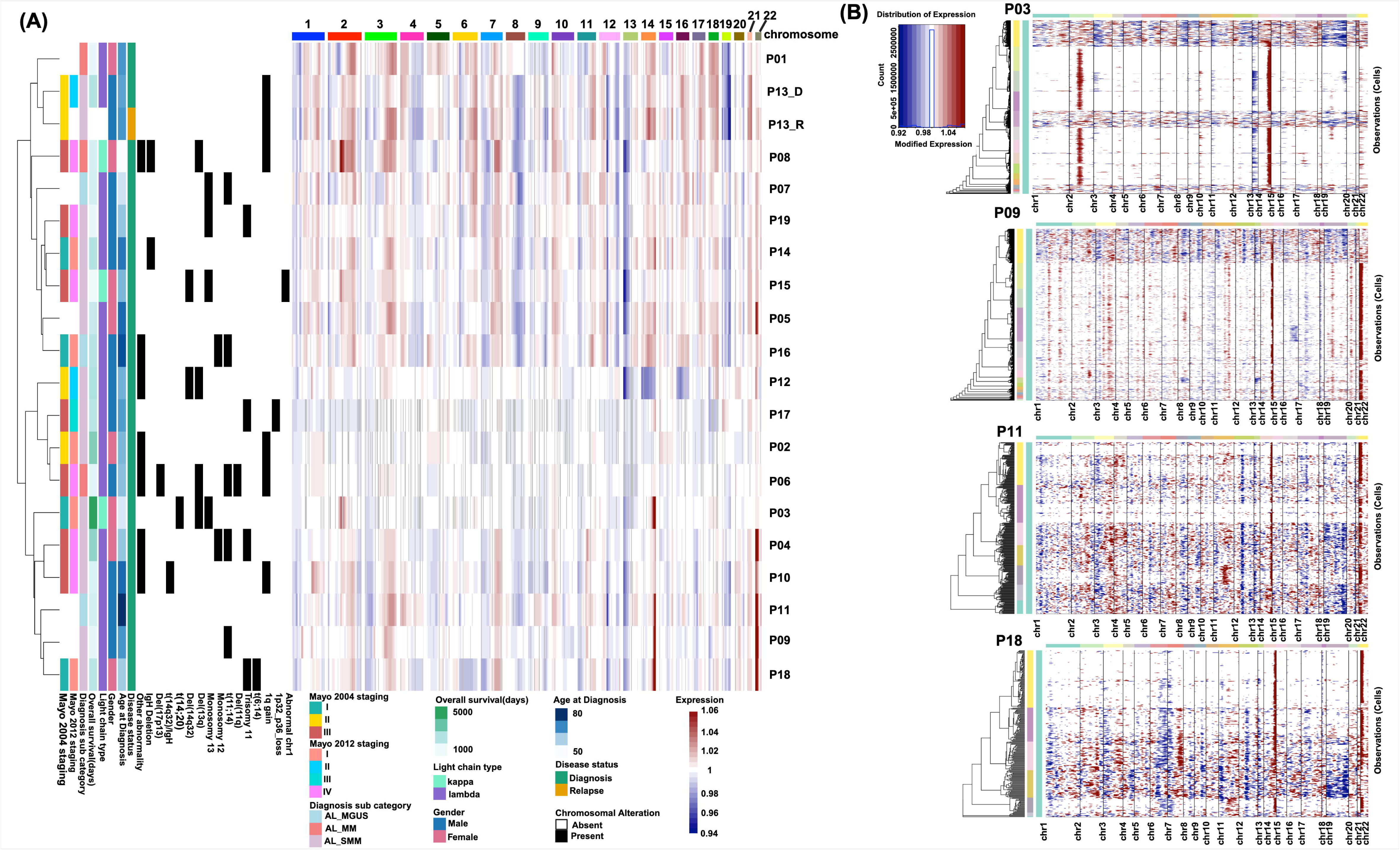
InferCNV Reveals Extensive Inter- and Intra-patient Heterogeneity in Plasma Cell Populations. **(A)** Heatmap showing inferCNV output, where gene expression was aggregated at the cytoband level by mapping each gene to its genomic cytoband; the left heatmap displays FISH-identified cytogenetic abnormalities, and the right heatmap shows cytoband-level copy-number variation across the cohort (n = 20). **(B)** InferCNV output from the four samples (P3, P9, P11 and P18) that showed the characteristic 14q gain pattern, with blue indicating deletions and red indicating amplifications.

Intra-patient heterogeneity was evident, with distinct PC subclusters displaying specific gains and losses, suggesting functionally divergent subpopulations within patients (**Supplementary Figure6**). Comparison with clinical FISH data showed partial concordance; inferCNV captured multiple large-scale alterations but had reduced sensitivity for focal or low-burden lesions consistent with previous reports^37^ (**Figure 6A**).

## DISCUSSION

Despite treatment advances, diagnosis of AL amyloidosis is often delayed, and early mortality remains high (≈13–20% within 6 months), especially in advanced cardiac stages^38^. Although genomic and transcriptional features have been described in AL amyloidosis, the cell-cell communication networks and immune-tumor interactions driving AL pathogenesis remain poorly understood^14,39^. Our single cell multiomic profiling provides an integrated framework for understanding how PCintrinsic programs, immune ecosystem remodelling, and genomic diversity collectively shape the biology of AL amyloidosis across its clinical continuum. By resolving >100,000 high quality cells into 13 immune and stromal compartments and six transcriptionally distinct PC states, our study reveals a coordinated pattern of tumor microenvironment interactions that has remained largely uncharacterized in AL. These data position AL not merely as a disorder of toxic light chain production, but as a complex ecosystem where malignant PCs rewire immune circuits, sculpt the BM niche, and evolve through both transcriptional and genomic diversification.

A key finding of our study is the inverse relationship between PC abundance and both CD4⁺ and CD8⁺ T-cell proportions, indicating that higher PC burden coincides with reduced T-cell presence. Samples enriched for CD4⁺ and CD8⁺ T cells also tended to present with lower Mayo stage, suggesting that preserved T-cell immunity aligns with less advanced disease. Single-cell profiling further showed the emergence of distinct exhaustion programs in CD4⁺, CD8⁺ and NK cell populations, highlighting an under recognized immune dysfunction axis in AL. Notably, both CD4⁺ T-cell exhaustion and NK cell *CCL3* expression exhibited prognostic associations, underscoring the clinical relevance of immune dysfunction even in a PC dominant malignancy. Few studies have assessed immune checkpoint expression in AL amyloidosis, with recent work reporting elevated expression of VISTA, PD1, and TIGIT on T cells in peripheral blood but limited to a small marker panel and bulk level analysis^40^. Further observation of *CCL3* expression in NK and CD4⁺ exhausted cells in AL is similar with findings in MM, where *CCL3* is enriched in GZMK TIGIT exhausted CD8⁺ T cells and CD4⁺ T-cell exhaustion, marked by CD4⁺PD-1⁺ subsets and marrow infiltrating regulatory T cells, correlates with inferior survival in patients with newly diagnosed MM^41,42^.

Our global cell-cell communication landscape in AL revealed several broad functional classes of signaling pathways: an adhesion and homing module (PECAM1/2, ICAM, SELPLG, SELL, CADM, NCAM, COLLAGEN, THBS, SEMA4/7A) mediating leukocyte-endothelium and PC stromal tethering; a chemokine and cytokine driven module (CCL, CXCL, MIF, MDK, galectins, GRN) shaping immune cell recruitment and tissue remodeling; a lipid and eicosanoid mediated immunometabolism module (cholesterol, prostaglandins, CysLTs, Annexin, CypA); an intracellular and damage associated signaling module (APP, CTSG, galectins, resistin, MDK) reflecting a stress and DAMP rich microenvironment; and T/NK centric modules (CD6, LCK, CD45, CD160, LAIR1, CLEC, MHC I/II, IFN II) captured activation, co-regulation, and antigen presentation contexts relevant to immune exhaustion. In our dataset, IFN II (IFNγ) was a prominent outgoing signal from lymphocyte subsets with strong exhaustion signatures, similarly reported in hematologic tumors where IFNγ secreting exhausted CD8^+^ T cells retain effector activity^43^. Within this network, CD14^+^ and CD16^+^ monocytes, together with GMPs and cDC2, act as dominant BAFF senders to B cells and PCs, establishing a myeloid-driven BAFF-BCMA survival niche. This axis represents a well-established pro-survival pathway in MM, now targeted by BCMA directed immunotherapies, with emerging evidence that BCMA and sBCMA also reflect disease activity in AL^44,45^.

PCs, although not the strongest overall interactors, send stress and migration related cues such as MDK, MIF, CTSG and SEMA7A, and receive multiple reinforcing survival signals including BAFF-BCMA, MIF:CD74-CXCR4 and PPIA:BSG, the latter two recently highlighted in MM as key axes between malignant PCs and their microenvironment^46^. Together, these interactions show that malignant PCs rely on layered support from surrounding immune and stromal populations while at the same time reshaping the BM niche into an increasingly immunosuppressive environment. In addition to receiving survival inputs, PCs activate several immune evasion mechanisms through MDK-NCL shaping an immunosuppressive microenvironment, MDK-VLA4 mediated adhesion, pan-HLA class I engagement of CD8^+^ T cells and HLA-E driven inhibition of NK cells v-ia CD94/NKG2A^47^. MDK-NCL is a well-supported signaling axis in solid tumors, and VLA-4 is a central adhesion integrin in hematologic malignancies^48,49^.

PC derived MDK, MIF, PPIA and PGE2 further remodel monocytes and GMPs driving them toward tumor supportive states, consistent with systemic monocytes ‘education’ seen across cancers^50^. This bidirectional communication is reinforced by feedback loops such as MIF:CD74-CXCR4, THBS1:CD47, and PECAM1 mediated adhesion, forming a tightly connected protection circuit between malignant PCs and myeloid cells. Prior work has implicated MIF-CD74-CXCR4 signaling and enhanced adhesion pathways as potential feedback axes between PCs and myeloid cells in MM^51^. Elevated *CXCR4* expression in GMPs and higher *PPIA* levels in CD16^+^ monocytes associated with poorer survival, highlighting these axes as both mechanistic drivers and potential biomarkers. Overall, recurrent MK, MIF, BAFF-BCMA, PECAM1/PECAM2 and CypA signaling links PCs with T cells, NK cells and myeloid populations in ways that enhance tumor cell persistence and suppress immune responses. These findings position microenvironment driven signaling, particularly MK and BAFF-BCMA pathways, as central to AL pathogenesis and highlight promising therapeutic targets.

Further, the study also revealed marked transcriptional heterogeneity among AL PCs, with subclusters differing in proliferation, survival, stress adaptation, and stromal interaction programs. Some populations displayed apoptosis resistant signatures characterized by *CCND1*, *MCL1*, *BCL2* and *MAFB*, while others showed stromal remodeling features marked by *SRGN* and *TPT1*, patterns that parallel observations in MM datasets^52,53^. Consistent with these signatures, recent studies demonstrate that AL PCs are highly apoptosis primed yet dependent on MCL1 and BCL2, and that BH3 mimetics can enhance responses to standard therapies^54^. Higher *IGHA1* expression correlated with poorer survival, suggesting that the transcriptional identity of amyloidogenic PCs contributes to clinical behavior and treatment sensitivity. Together, these findings support a model in which AL PCs occupy multiple malignant states that differentially influence amyloid production, microenvironment remodeling, and therapy response.

We also demonstrated a clear transcriptional divergence between κ and λ AL plasma cell subtypes. λ AL is characterized by inflammatory, TNF/NFκB, interferon-γ, apoptotic, and EMT-like pathways, whereas λ AL exhibits MYC-driven metabolic and ER stress programs with lower inflammatory signaling. These subtype-specific architectures provide a molecular basis for clinical observations that λ AL is more prevalent but κ AL often shows distinct organ involvement and survival dynamics ^5,55,56^. Our findings introduce a mechanistic rationale for these differences and suggest that κ and λ AL may benefit from subtype specific therapeutic strategies targeting inflammatory versus metabolic vulnerabilities.

Across precursor states, our analysis identifies a coherent MYC-p53-apoptosis axis that differentiates AL_MGUS, AL_SMM, and AL with concomitant MM (AL_MM). While PC from these categories intermingle transcriptomically, differential expression and pathway analysis revealed a directional shift toward stress survival signaling and suppression of interferon mediated immune programs in more advanced disease. These findings suggest that progression in AL is not defined by wholesale transcriptomic transformation but rather by selective amplification of stress adaptation pathways and loss of immune reactivity, offering a new conceptual model of clonal evolution in AL distinct from the classical MGUS-SMM-MM paradigm^57^.

Finally, our inferCNV^27^ analysis revealed widespread structural variation, including recurrent 1q gains, patient specific CNVs, and marked inter- and intra-patient heterogeneity. We also detected additional amplifications and deletions not captured by FISH, many of which appeared consistently across multiple samples, highlighting that inferCNV adds novelty by uncovering CNVs beyond standard FISH and underscoring the genetic complexity of AL PCs in this low purity, microenvironment dominant setting.

Together, these data establish a systems level blueprint of AL amyloidosis, integrating immune dysfunction, plasma cell heterogeneity, signaling crosstalk, light chain subtype biology, and genomic architecture. This framework provides mechanistic insight into AL pathogenesis, refines the biological basis of clinical subgroups, and reveals multiple candidate pathways including T-cell/NK exhaustion circuits, MK/MIF centered interactions, λ UPR *vs.* κ inflammatory programs, and MYC-p53-apoptosis progression signals that warrant exploration as therapeutic targets. Future work leveraging larger cohorts and paired longitudinal sampling will be essential to validate prognostic markers and determine how immune and plasma cell ecotypes evolve in response to therapy.

## Data Availability

All data produced in the present study are available upon reasonable request to the authors

## ACKNOWLEDGEMENTS

The authors are grateful for the generous donation of samples from the contributing patients. The samples were obtained from the Finnish Hematology Registry and Biobank. The authors also thank the FIMM Single-Cell Analytics and Genomics Units, which are supported by HiLIFE and Biocenter Finland. This work has been supported by funding from Oncopeptides, Sigrid Jusélius Foundation, Cancer Foundation Finland, Research Council of Finland (No. 334781, 320185, 352265 and 357686). We acknowledge the use of ChatGPT for text formatting and language polishing. The authors also acknowledge the use of BioRender.com for the generation of figure illustrations.

## AUTHORSHIP CONTRIBUTIONS

CAH, AS, and RK contributed to the conception/design of the study with support from KA, SL. Data analysis was performed by AS with advice from RK and CAH. AS and RK wrote the manuscript with support from CAH. JJM and MHS contributed to single cell sequencing sample preparation. SL provided clinical data related to the samples. All authors critically reviewed and contributed to the final manuscript.

