## Supplementary Figures for "Cellular Diversity, Immune Crosstalk, and Genomic Alterations in Light Chain Amyloidosis"

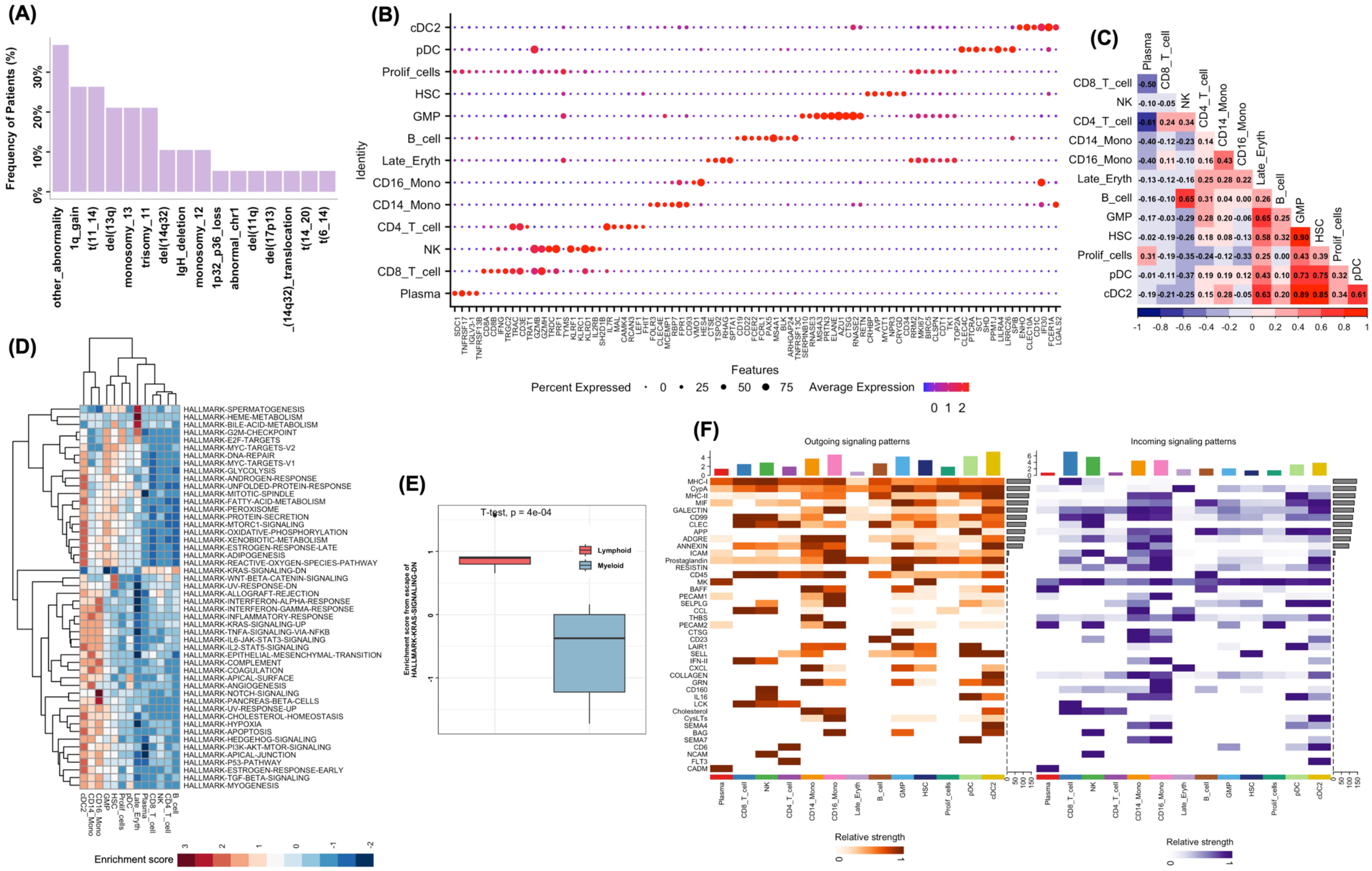


**Supplementary Figure1.** (A) The frequency of the cytogenetic aberrations identified through fluorescent in situ hybridization (FISH) in our AL study cohort. (B) A dot plot generated using Seurat, displaying the top 5 marker genes used for cell type annotation. (C) Correlation of the proportions of various predicted cell types across 20 samples. (D) Pathway enrichment analysis of different cell types. (E) Boxplot showing the difference in the KRAS signaling- DN in lymphoid and myeloid cells. (F) Outgoing and Incoming Signaling Patterns Across 41 Pathways in the Amyloidosis Landscape.

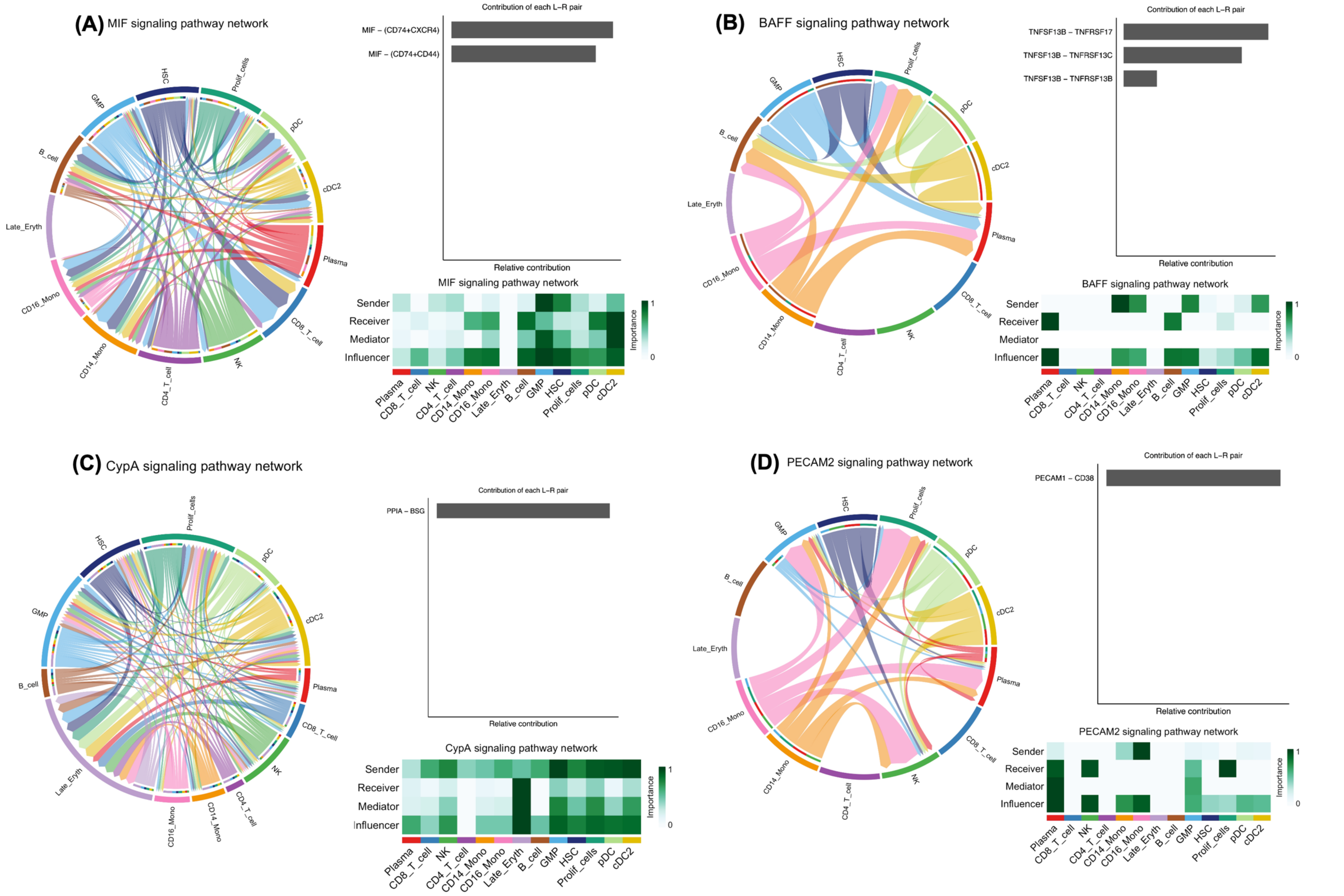
(A) MIF signaling pathway network
Contribution of each L–R pair
MIF – (CD74+CXCR4)
MIF – (CD74+CD44)
Relative contribution
MIF signaling pathway network
Sender
Receiver
Mediator
Influencer
Importance
(B) BAFF signaling pathway network
Contribution of each L–R pair
TNFSF13B – TNFRSF17
TNFSF13B – TNFRSF13C
TNFSF13B – TNFRSF13B
Relative contribution
BAFF signaling pathway network
(C) CypA signaling pathway network
Contribution of each L–R pair
PPIA – BSG
Relative contribution
CypA signaling pathway network
(D) PECAM2 signaling pathway network
Contribution of each L–R pair
PECAM1 – CD38
Relative contribution
PECAM2 signaling pathway network
Plasma
CD8_T_cell
NK
CD4_T_cell
CD14_Mono
CD16_Mono
Late_Eryth
B_cell
GMP
HSC
Prolif_cells
pDC
cDC2

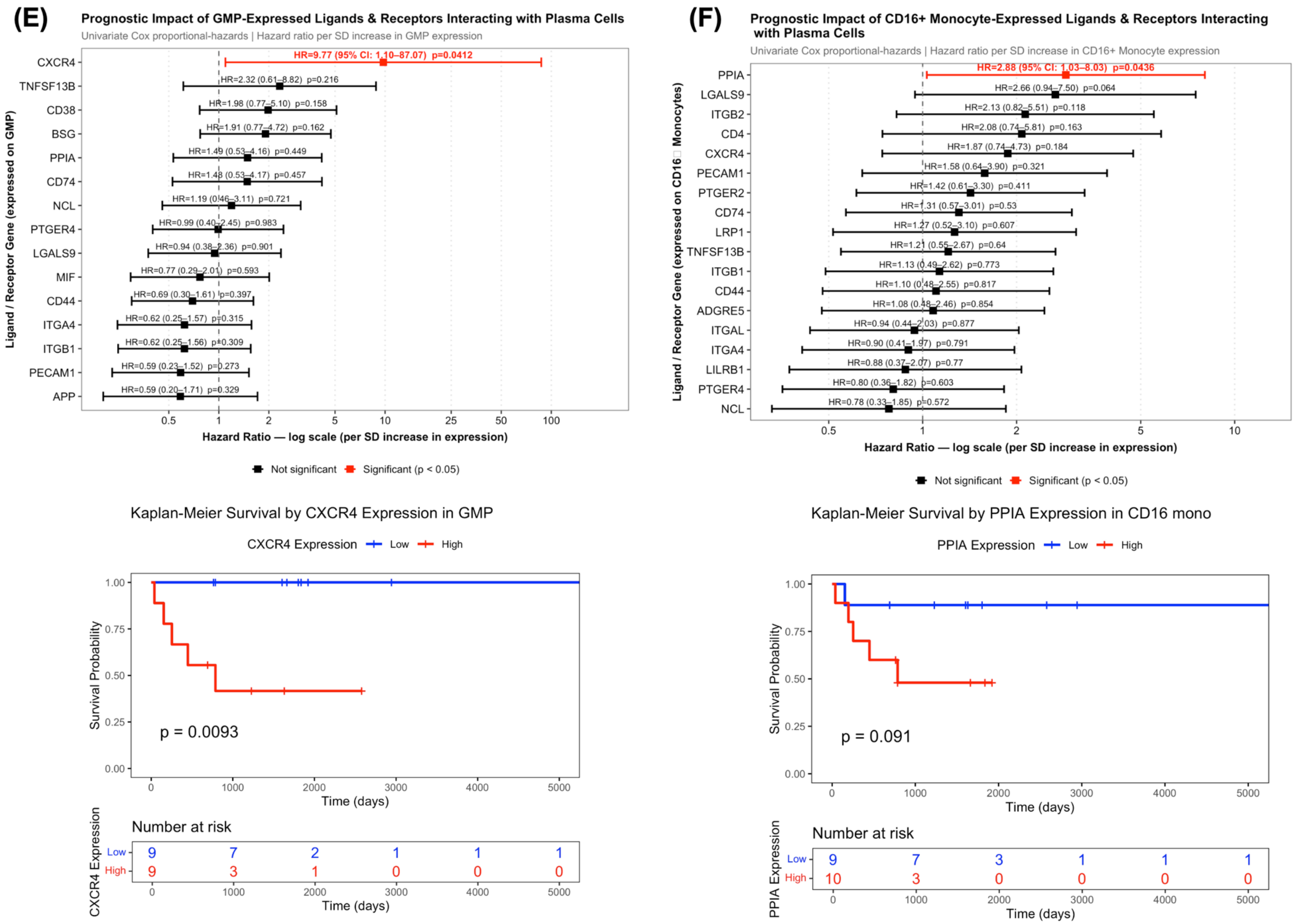


**Supplementary Figure2.** Figure 2A–D summarizes four major signaling pathways using chord diagrams to depict communication directionality across cell types, bar plots to show the contribution of each ligand–receptor pair, and heatmaps to identify dominant senders, receivers, mediators, and influencers within each network. (A) MIF pathway (B) BAFF pathway (C) CypA pathway (D) PECAM2 pathway. (E) Prognostic impact of GMP-expressed ligands and receptors interacting with plasma cells in amyloidosis. (E – lower panel) Kaplan–Meier survival analysis based on median stratification of CXCR4 expression in GMP Cells. (F) Prognostic impact of $CD16^{+}$ monocyte-expressed ligands and receptors interacting with plasma cells. (F – lower panel) Kaplan–Meier survival analysis based on median stratification of PPIA expression in $CD16^{+}$ monocyte.

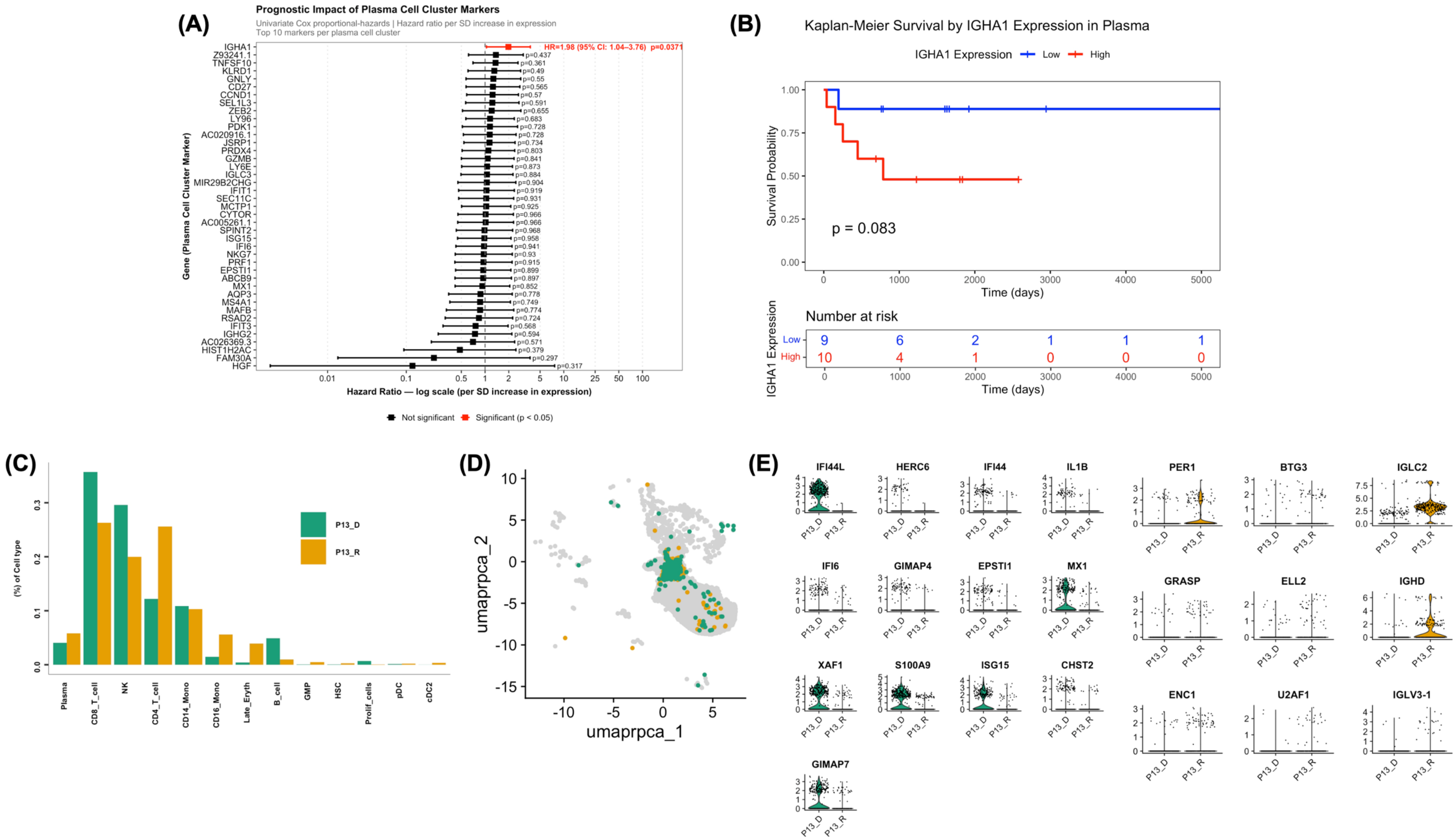


**Supplementary Figure3.** (A) A forest plot summarizing the prognostic impact of plasma-cell–expressed markers, displaying hazard ratios and confidence intervals to identify genes associated with favorable or unfavorable clinical outcomes. (B) A Kaplan–Meier curve based on median stratification of IGH4 expression in plasma cells, illustrating differences in survival probability between the high- and low-expression groups. (C) Comparison of the proportions of major cell types in patient P13 at both the diagnosis and relapse stages. (D) UMAP visualization showing the distribution and clustering of plasma cells from patient P13 at diagnosis and relapse. (E) Boxplots illustrating differentially expressed genes in plasma cells between the diagnosis and relapse stages for patient P13.

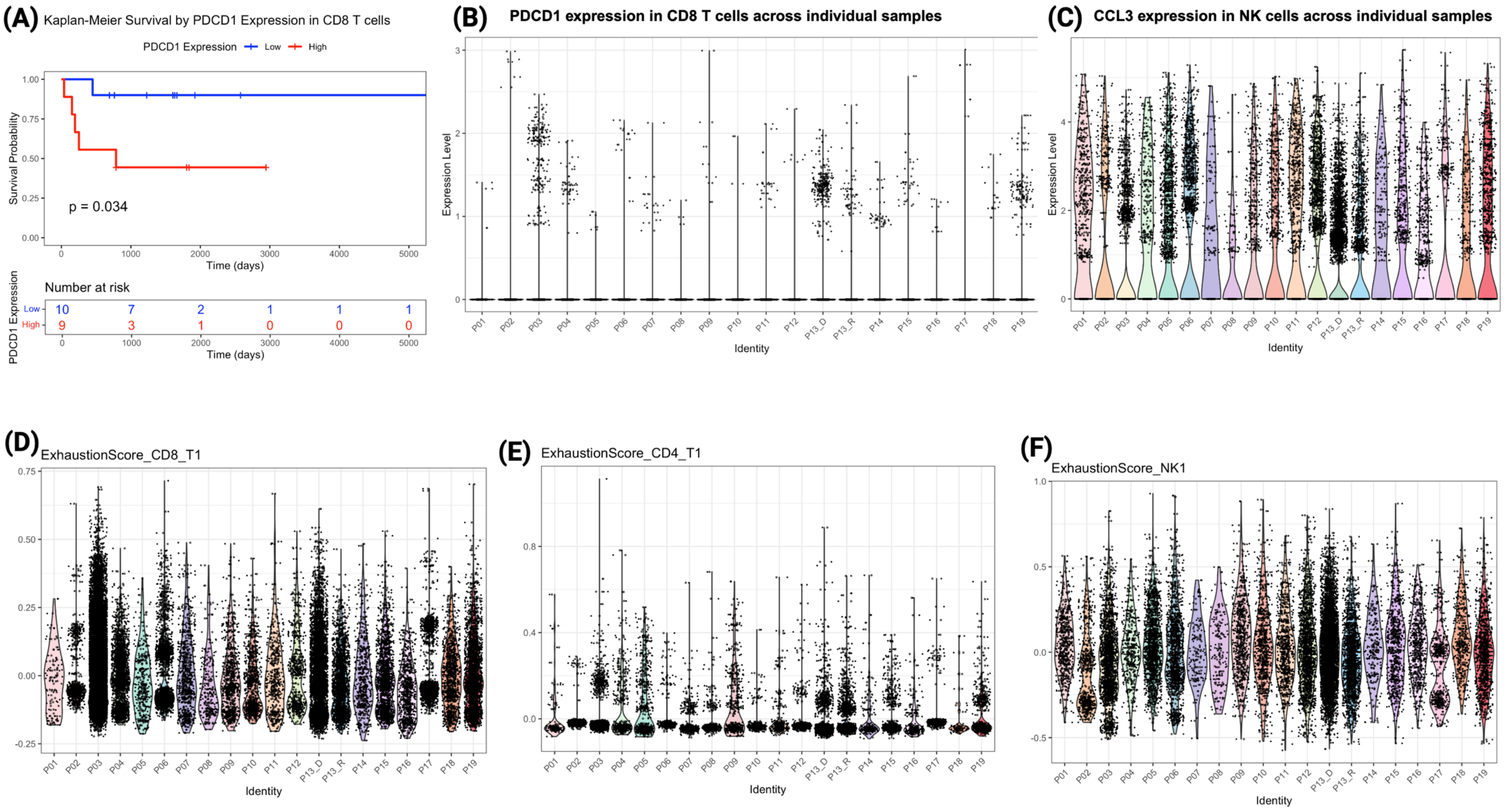


**Supplementary Figure4.** (A) Kaplan–Meier curve based on median stratification of PDCD1 expression in CD8 T cells, showing reduced survival in the high expression group (log rank p = 0.034), an effect not observed in the Cox proportional hazards analysis. (B) PDCD1 expression levels in CD8 T cells across individual samples. (C) CCL3 expression levels in NK cells across individual samples. (D) Exhaustion score distribution in CD8 T cells. (E) Exhaustion score distribution in CD4 T cells. (F) Exhaustion score distribution in NK cells.

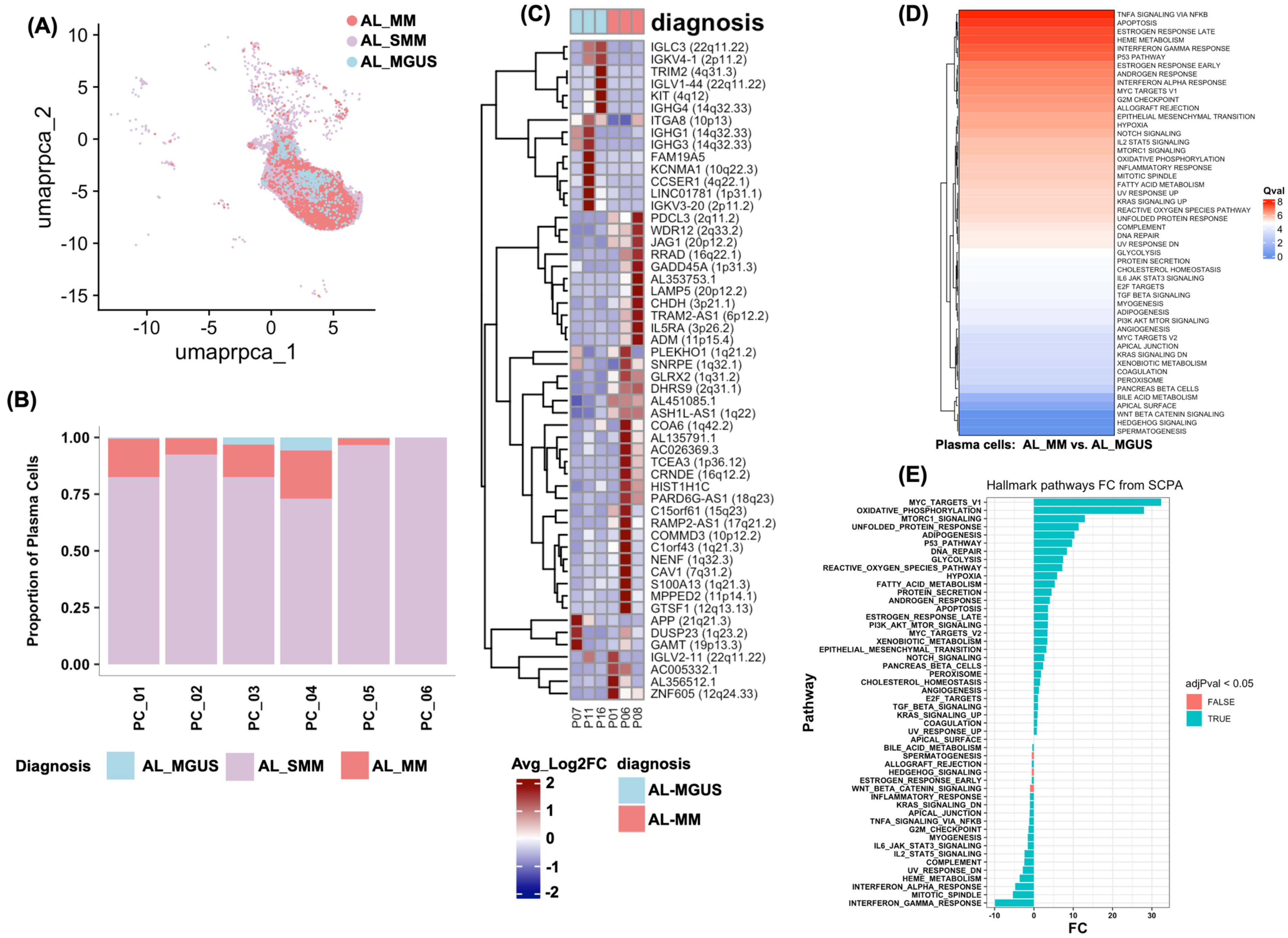

(A)
AL_MM
AL_SMM
AL_MGUS
umaprpca_2
umaprpca_1
(B)
Proportion of Plasma Cells
PC_01
PC_02
PC_03
PC_04
PC_05
PC_06
Diagnosis AL_MGUS AL_SMM AL_MM
(C)
diagnosis
IGLC3 (22q11.22)
IGKV4-1 (2p11.2)
TRIM2 (4q31.3)
IGLV1-44 (22q11.22)
KIT (4q12)
IGHG4 (14q32.33)
ITGA8 (10p13)
IGHG1 (14q32.33)
IGHG3 (14q32.33)
FAM19A5
KCNMA1 (10q22.3)
CCSER1 (4q22.1)
LINC01781 (1p31.1)
IGKV3-20 (2p11.2)
PDCL3 (2q11.2)
WDR12 (2q33.2)
JAG1 (20p12.2)
RRAD (16q22.1)
GADD45A (1p31.3)
AL353753.1
LAMP5 (20p12.2)
CHDH (3p21.1)
TRAM2-AS1 (6p12.2)
IL5RA (3p26.2)
ADM (11p15.4)
PLEKHO1 (1q21.2)
SNRPE (1q32.1)
GLRX2 (1q31.2)
DHRS9 (2q31.1)
AL451085.1
ASH1L-AS1 (1q22)
COA6 (1q42.2)
AL135791.1
AC026369.3
TCEA3 (1p36.12)
CRNDE (16q12.2)
HIST1H1C
PARD6G-AS1 (18q23)
C15orf61 (15q23)
RAMP2-AS1 (17q21.2)
COMMD3 (10p12.2)
C1orf43 (1q21.3)
NENF (1q32.3)
CAV1 (7q31.2)
S100A13 (1q21.3)
MPPED2 (11p14.1)
GTSF1 (12q13.13)
APP (21q21.3)
DUSP23 (1q23.2)
GAMT (19p13.3)
IGLV2-11 (22q11.22)
AC005332.1
AL356512.1
ZNF605 (12q24.33)
P07 P11 P16 P01 P06 P08
Avg_Log2FC
diagnosis
AL-MGUS
AL-MM
(D)
TNFA SIGNALING VIA NFKB
APOPTOSIS
ESTROGEN RESPONSE LATE
HEME METABOLISM
INTERFERON GAMMA RESPONSE
P53 PATHWAY
ESTROGEN RESPONSE EARLY
ANDROGEN RESPONSE
INTERFERON ALPHA RESPONSE
MYC TARGETS V1
G2M CHECKPOINT
ALLOGRAFT REJECTION
EPITHELIAL MESENCHYMAL TRANSITION
HYPOXIA
NOTCH SIGNALING
IL2 STAT5 SIGNALING
MTORC1 SIGNALING
OXIDATIVE PHOSPHORYLATION
INFLAMMATORY RESPONSE
MITOTIC SPINDLE
FATTY ACID METABOLISM
UV RESPONSE UP
KRAS SIGNALING UP
REACTIVE OXYGEN SPECIES PATHWAY
UNFOLDED PROTEIN RESPONSE
COMPLEMENT
DNA REPAIR
UV RESPONSE DN
GLYCOLYSIS
PROTEIN SECRETION
CHOLESTEROL HOMEOSTASIS
IL6 JAK STAT3 SIGNALING
E2F TARGETS
TGF BETA SIGNALING
MYOGENESIS
ADIPOGENESIS
PI3K AKT MTOR SIGNALING
ANGIOGENESIS
MYC TARGETS V2
APICAL JUNCTION
KRAS SIGNALING DN
XENOBIOTIC METABOLISM
COAGULATION
PEROXISOME
PANCREAS BETA CELLS
BILE ACID METABOLISM
APICAL SURFACE
WNT BETA CATENIN SIGNALING
HEDGEHOG SIGNALING
SPERMATOGENESIS
Qval
Plasma cells: AL_MM vs. AL_MGUS
(E)
Hallmark pathways FC from SCPA
Pathway
FC
adjPval < 0.05
FALSE
TRUE

(F)

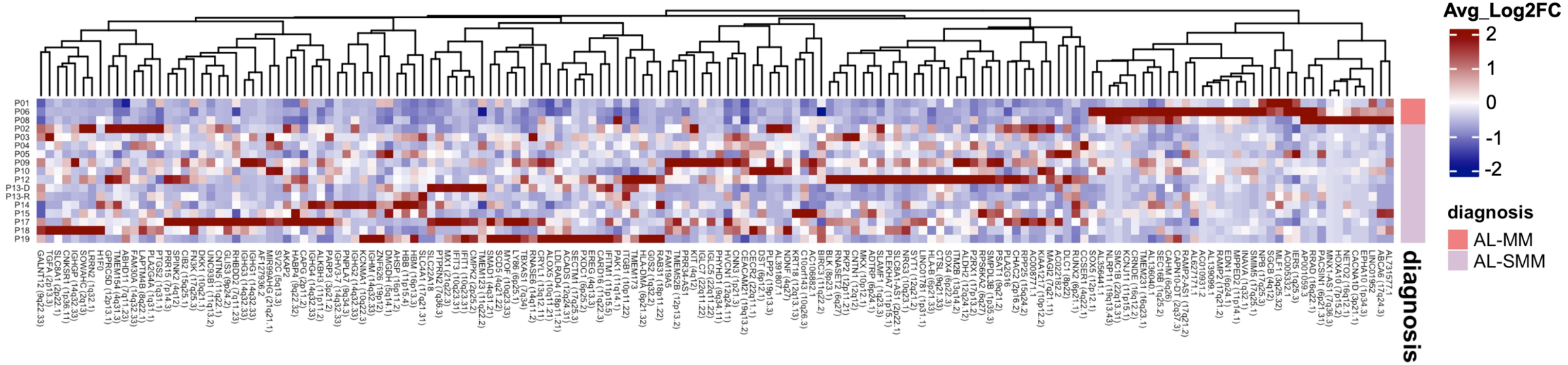

Avg_Log2FC
2
1
0
-1
-2
diagnosis
AL-MM
AL-SMM
P01
P06
P08
P02
P03
P04
P05
P09
P10
P12
P13-D
P13-R
P14
P15
P17
P18
P19
diagnosis


(G)

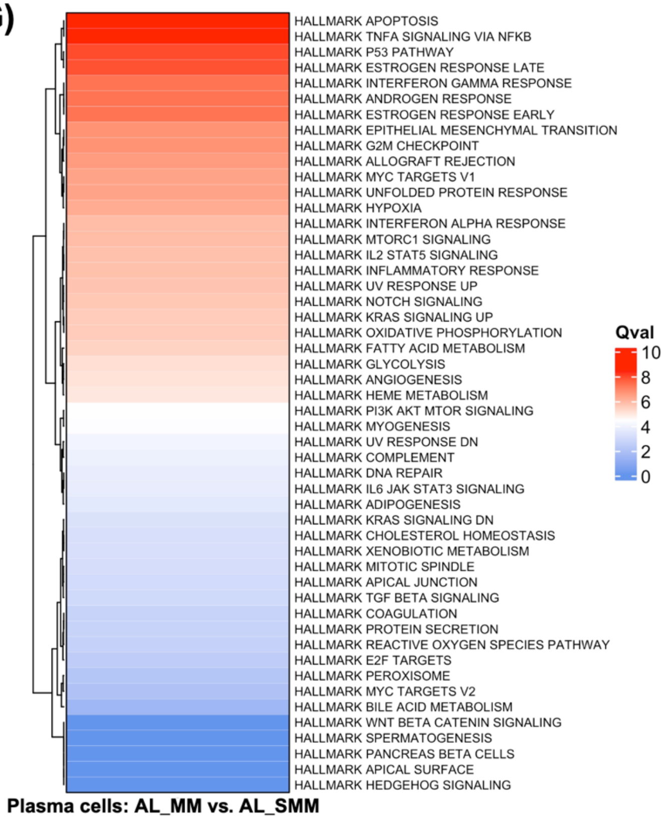

HALLMARK APOPTOSIS
HALLMARK TNFA SIGNALING VIA NFKB
HALLMARK P53 PATHWAY
HALLMARK ESTROGEN RESPONSE LATE
HALLMARK INTERFERON GAMMA RESPONSE
HALLMARK ANDROGEN RESPONSE
HALLMARK ESTROGEN RESPONSE EARLY
HALLMARK EPITHELIAL MESENCHYMAL TRANSITION
HALLMARK G2M CHECKPOINT
HALLMARK ALLOGRAFT REJECTION
HALLMARK MYC TARGETS V1
HALLMARK UNFOLDED PROTEIN RESPONSE
HALLMARK HYPOXIA
HALLMARK INTERFERON ALPHA RESPONSE
HALLMARK MTORC1 SIGNALING
HALLMARK IL2 STAT5 SIGNALING
HALLMARK INFLAMMATORY RESPONSE
HALLMARK UV RESPONSE UP
HALLMARK NOTCH SIGNALING
HALLMARK KRAS SIGNALING UP
HALLMARK OXIDATIVE PHOSPHORYLATION
HALLMARK FATTY ACID METABOLISM
HALLMARK GLYCOLYSIS
HALLMARK ANGIOGENESIS
HALLMARK HEME METABOLISM
HALLMARK PI3K AKT MTOR SIGNALING
HALLMARK MYOGENESIS
HALLMARK UV RESPONSE DN
HALLMARK COMPLEMENT
HALLMARK DNA REPAIR
HALLMARK IL6 JAK STAT3 SIGNALING
HALLMARK ADIPOGENESIS
HALLMARK KRAS SIGNALING DN
HALLMARK CHOLESTEROL HOMEOSTASIS
HALLMARK XENOBIOTIC METABOLISM
HALLMARK MITOTIC SPINDLE
HALLMARK APICAL JUNCTION
HALLMARK TGF BETA SIGNALING
HALLMARK COAGULATION
HALLMARK PROTEIN SECRETION
HALLMARK REACTIVE OXYGEN SPECIES PATHWAY
HALLMARK E2F TARGETS
HALLMARK PEROXISOME
HALLMARK MYC TARGETS V2
HALLMARK BILE ACID METABOLISM
HALLMARK WNT BETA CATENIN SIGNALING
HALLMARK SPERMATOGENESIS
HALLMARK PANCREAS BETA CELLS
HALLMARK APICAL SURFACE
HALLMARK HEDGEHOG SIGNALING
Qval
10
8
6
4
2
0
Plasma cells: AL_MM vs. AL_SMM


(H)

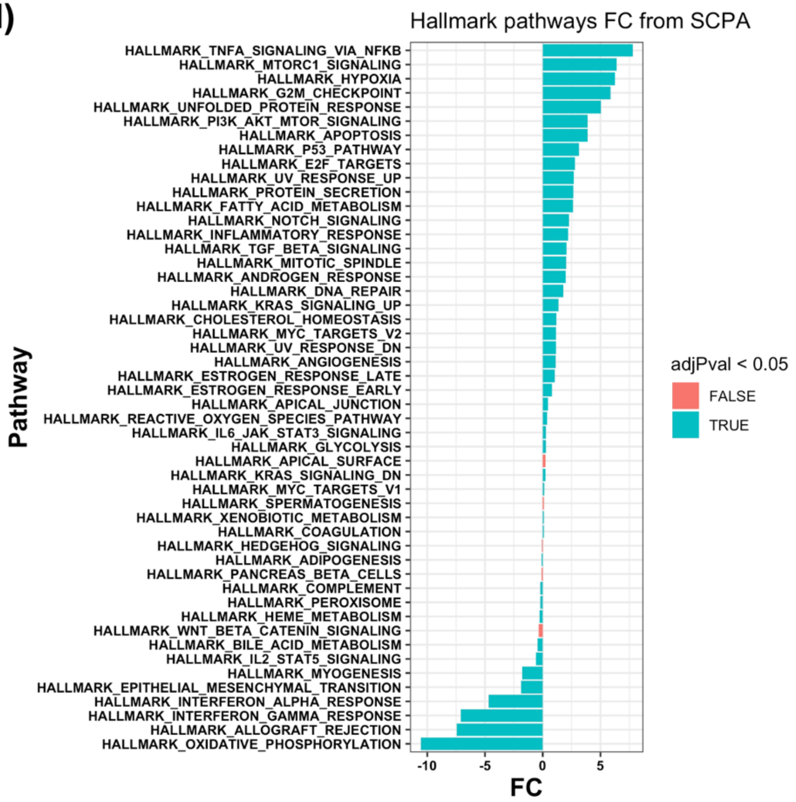

Hallmark pathways FC from SCPA
Pathway
HALLMARK_TNFA_SIGNALING_VIA_NFKB
HALLMARK_MTORC1_SIGNALING
HALLMARK_HYPOXIA
HALLMARK_G2M_CHECKPOINT
HALLMARK_UNFOLDED_PROTEIN_RESPONSE
HALLMARK_PI3K_AKT_MTOR_SIGNALING
HALLMARK_APOPTOSIS
HALLMARK_P53_PATHWAY
HALLMARK_E2F_TARGETS
HALLMARK_UV_RESPONSE_UP
HALLMARK_PROTEIN_SECRETION
HALLMARK_FATTY_ACID_METABOLISM
HALLMARK_NOTCH_SIGNALING
HALLMARK_INFLAMMATORY_RESPONSE
HALLMARK_TGF_BETA_SIGNALING
HALLMARK_MITOTIC_SPINDLE
HALLMARK_ANDROGEN_RESPONSE
HALLMARK_DNA_REPAIR
HALLMARK_KRAS_SIGNALING_UP
HALLMARK_CHOLESTEROL_HOMEOSTASIS
HALLMARK_MYC_TARGETS_V2
HALLMARK_UV_RESPONSE_DN
HALLMARK_ANGIOGENESIS
HALLMARK_ESTROGEN_RESPONSE_LATE
HALLMARK_ESTROGEN_RESPONSE_EARLY
HALLMARK_APICAL_JUNCTION
HALLMARK_REACTIVE_OXYGEN_SPECIES_PATHWAY
HALLMARK_IL6_JAK_STAT3_SIGNALING
HALLMARK_GLYCOLYSIS
HALLMARK_APICAL_SURFACE
HALLMARK_KRAS_SIGNALING_DN
HALLMARK_MYC_TARGETS_V1
HALLMARK_SPERMATOGENESIS
HALLMARK_XENOBIOTIC_METABOLISM
HALLMARK_COAGULATION
HALLMARK_HEDGEHOG_SIGNALING
HALLMARK_ADIPOGENESIS
HALLMARK_PANCREAS_BETA_CELLS
HALLMARK_COMPLEMENT
HALLMARK_PEROXISOME
HALLMARK_HEME_METABOLISM
HALLMARK_WNT_BETA_CATENIN_SIGNALING
HALLMARK_BILE_ACID_METABOLISM
HALLMARK_IL2_STAT5_SIGNALING
HALLMARK_MYOGENESIS
HALLMARK_EPITHELIAL_MESENCHYMAL_TRANSITION
HALLMARK_INTERFERON_ALPHA_RESPONSE
HALLMARK_INTERFERON_GAMMA_RESPONSE
HALLMARK_ALLOGRAFT_REJECTION
HALLMARK_OXIDATIVE_PHOSPHORYLATION
-10
-5
0
5
FC
adjPval < 0.05
FALSE
TRUE

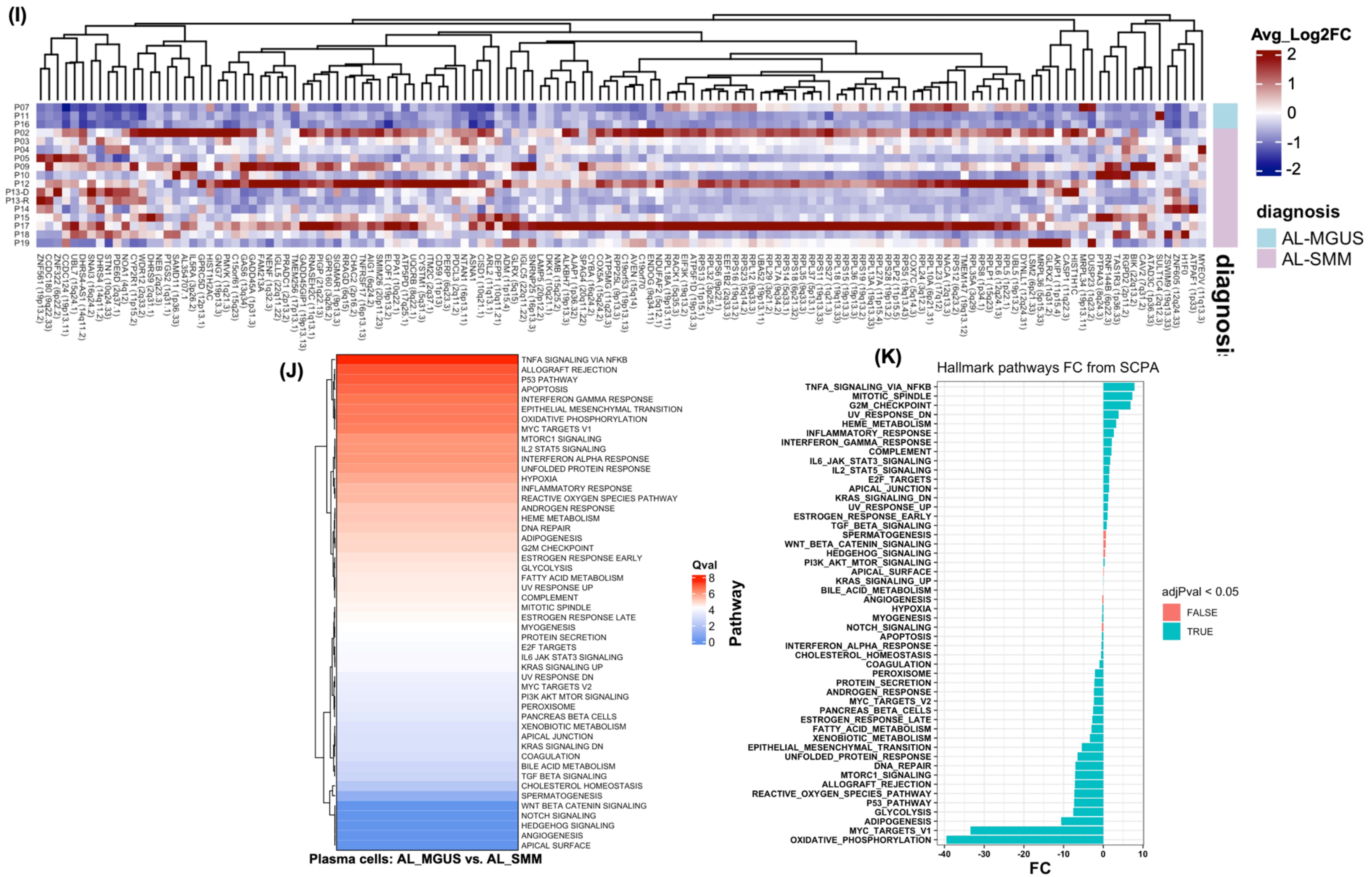


**Supplementary Figure5.** (A) UMAP projection of plasma cells annotated by disease stage, distinguishing AL_MM, AL_SMM, and AL_MGUS, highlighting transcriptional separation and overlap and stage-associated clustering patterns. (B) Proportion of plasma cells from AL_MM, AL_SMM, and AL_MGUS across each plasma cell cluster, illustrating shifts in cellular composition across disease progression. (C) Heatmap showing differentially regulated genes between AL_MM and AL_MGUS plasma cells. (D) Relative pathway enrichment analysis comparing AL_MM with AL_MGUS using Single-Cell Pathway Analysis (SCPA), where q-values quantify the degree of pathway divergence, with higher q-values indicating more pronounced differences. (E) Diverging bar chart displaying fold-change (FC) enrichment scores for pathways differentially enriched between AL_MM and AL_MGUS plasma cell populations. (F) Heatmap showing the differentially regulated genes between AL_MM and AL_SMM plasma cells. (G) Relative pathway enrichment analysis comparing AL_MM with AL_SMM using SCPA. (H) Diverging bar chart displaying fold-change (FC) enrichment scores for pathways differentially enriched between AL_MM and AL_SMM plasma cell populations. (I) Heatmap showing the differentially regulated genes between AL_MGUS and AL_SMM plasma cells. (J) Relative pathway enrichment analysis comparing AL_MGUS with AL_SMM using Single Cell Pathway Analysis (SCPA) (K) Diverging bar chart displaying fold-change (FC) enrichment scores for pathways differentially enriched between AL_MGUS and AL_SMM plasma cell populations.

P01, n= 99

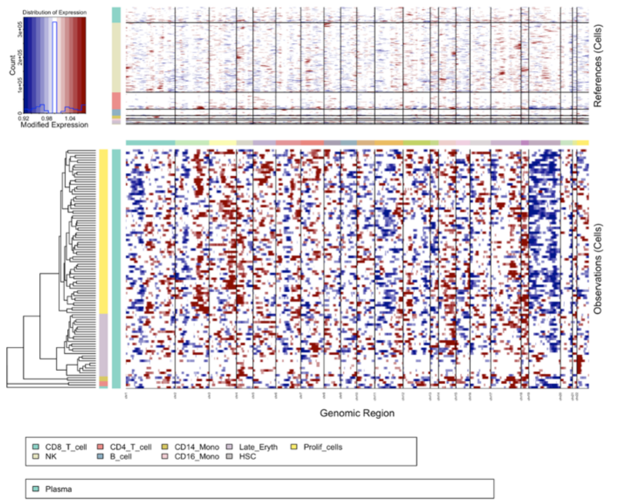


P02, n= 11019

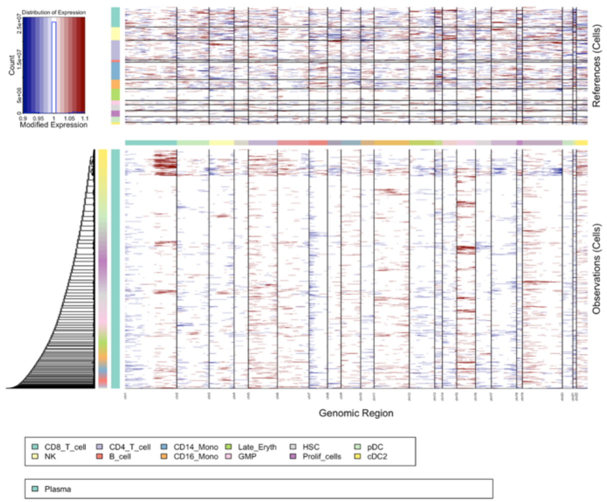


P03, n= 1073

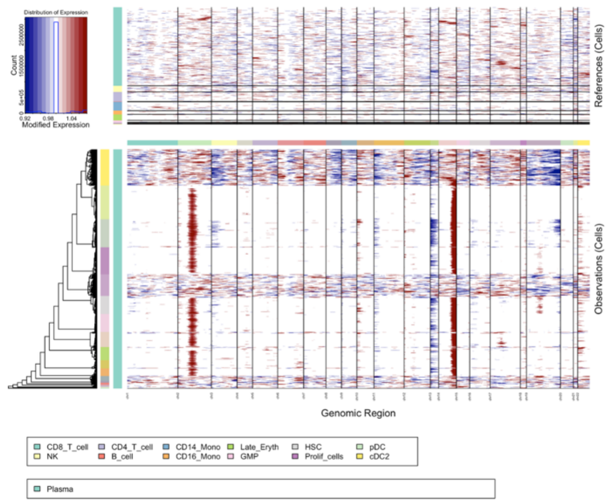


P04, n= 461

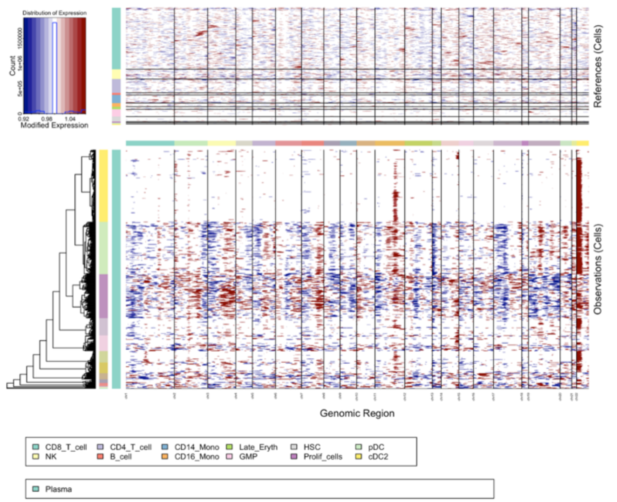


P05, n= 132

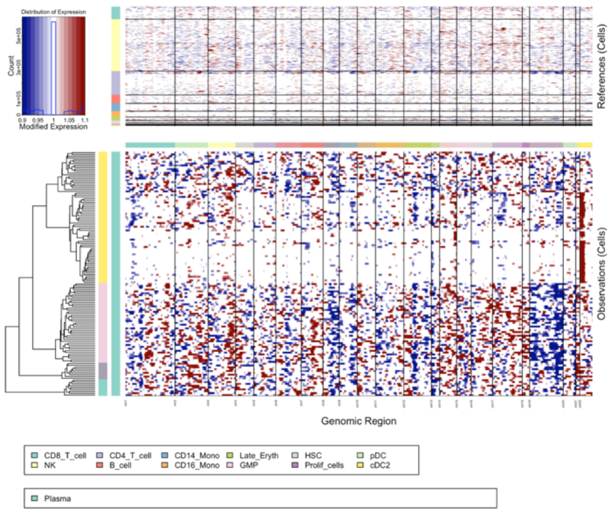


P06, n= 3947

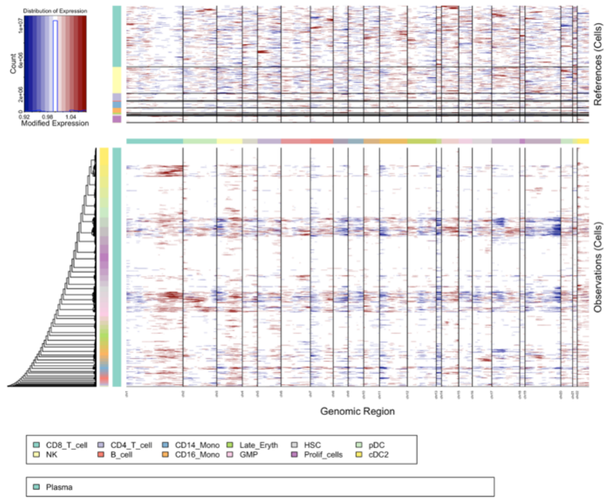

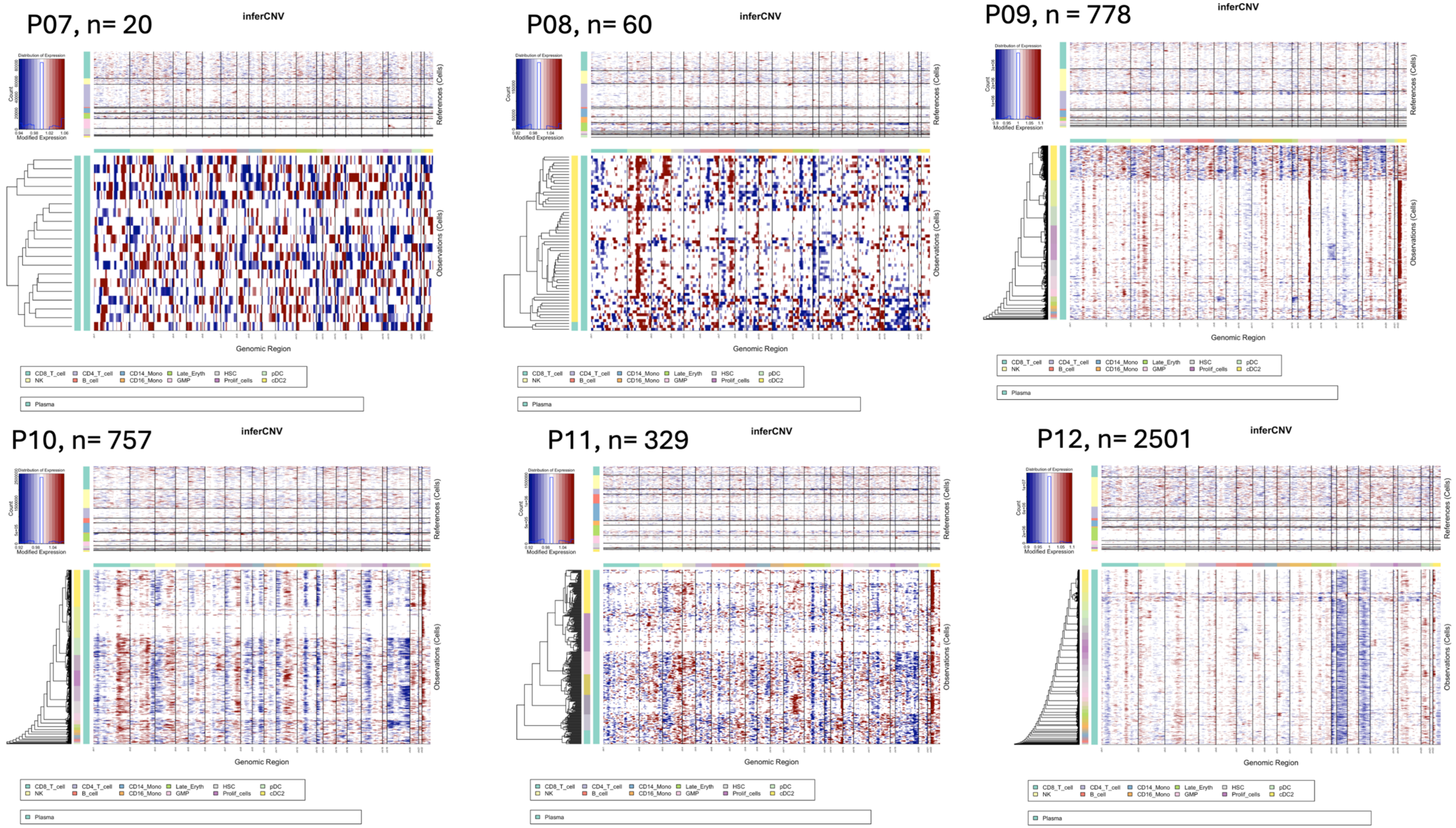

P07, n= 20
P08, n= 60
P09, n = 778
P10, n= 757
P11, n= 329
P12, n= 2501
inferCNV
Distribution of Expression
Count
Modified Expression
References (Cells)
Observations (Cells)
Genomic Region
CD8_T_cell
NK
CD4_T_cell
B_cell
CD14_Mono
CD16_Mono
Late_Eryth
GMP
HSC
Prolif_cells
pDC
cDC2
Plasma

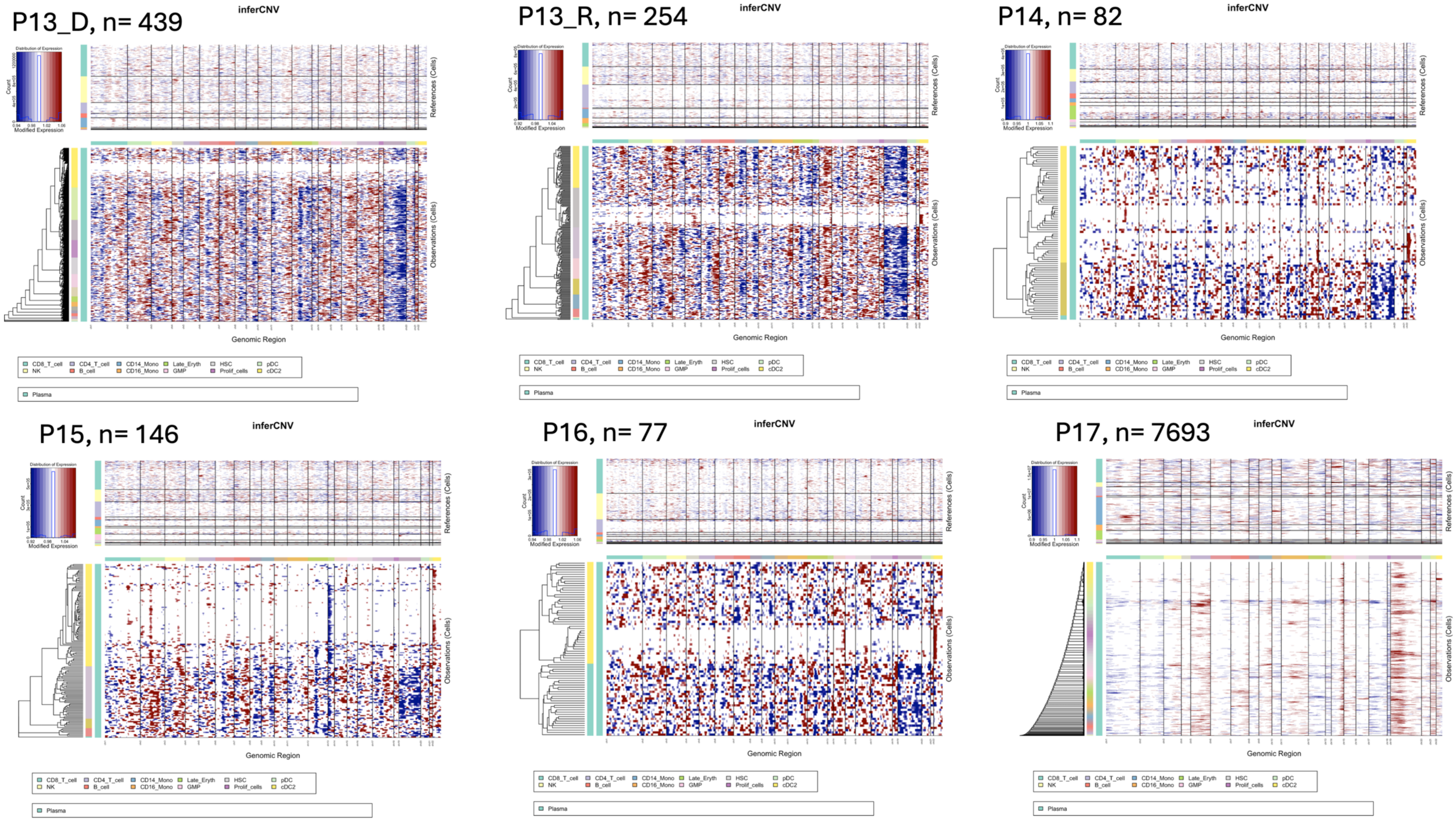

P13_D, n= 439
inferCNV
Distribution of Expression
Count
Modified Expression
References (Cells)
Observations (Cells)
Genomic Region
CD8_T_cell
NK
CD4_T_cell
B_cell
CD14_Mono
CD16_Mono
Late_Eryth
GMP
HSC
Prolif_cells
pDC
cDC2
Plasma
P13_R, n= 254
inferCNV
Distribution of Expression
Count
Modified Expression
References (Cells)
Observations (Cells)
Genomic Region
CD8_T_cell
NK
CD4_T_cell
B_cell
CD14_Mono
CD16_Mono
Late_Eryth
GMP
HSC
Prolif_cells
pDC
cDC2
Plasma
P14, n= 82
inferCNV
Distribution of Expression
Count
Modified Expression
References (Cells)
Observations (Cells)
Genomic Region
CD8_T_cell
NK
CD4_T_cell
B_cell
CD14_Mono
CD16_Mono
Late_Eryth
GMP
HSC
Prolif_cells
pDC
cDC2
Plasma
P15, n= 146
inferCNV
Distribution of Expression
Count
Modified Expression
References (Cells)
Observations (Cells)
Genomic Region
CD8_T_cell
NK
CD4_T_cell
B_cell
CD14_Mono
CD16_Mono
Late_Eryth
GMP
HSC
Prolif_cells
pDC
cDC2
Plasma
P16, n= 77
inferCNV
Distribution of Expression
Count
Modified Expression
References (Cells)
Observations (Cells)
Genomic Region
CD8_T_cell
NK
CD4_T_cell
B_cell
CD14_Mono
CD16_Mono
Late_Eryth
GMP
HSC
Prolif_cells
pDC
cDC2
Plasma
P17, n= 7693
inferCNV
Distribution of Expression
Count
Modified Expression
References (Cells)
Observations (Cells)
Genomic Region
CD8_T_cell
NK
CD4_T_cell
B_cell
CD14_Mono
CD16_Mono
Late_Eryth
GMP
HSC
Prolif_cells
pDC
cDC2
Plasma

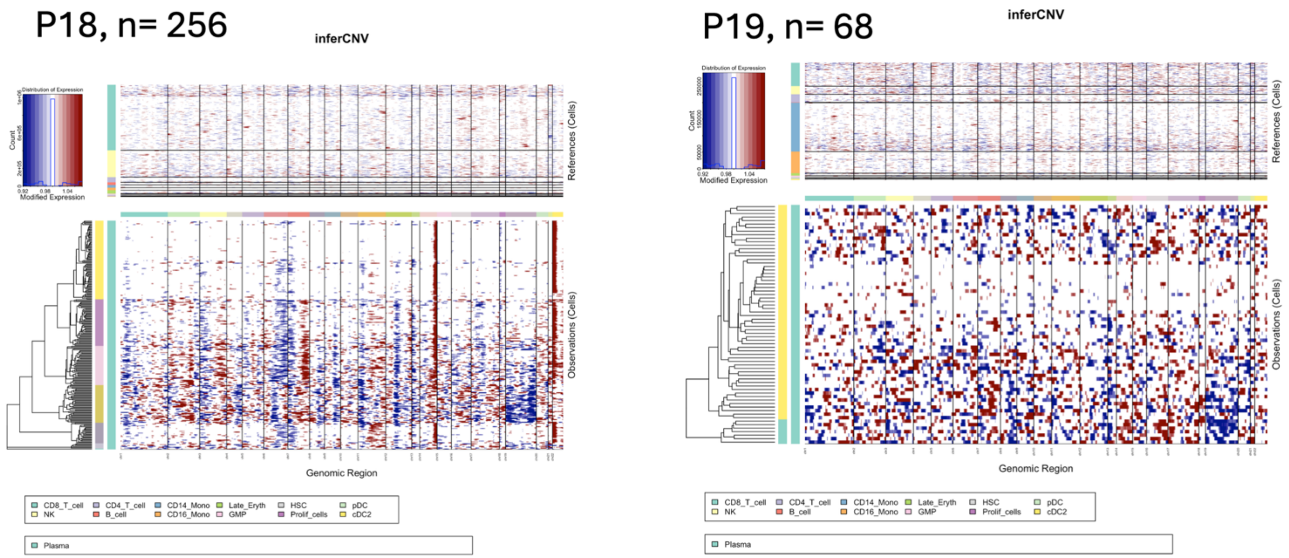


**Supplementary Figure6.** This figure represents the output of InferCNV, displaying inferred copy number variations (CNVs) across single cells. Each row corresponds to an individual cell while each column represents a genomic position ordered by chromosomal location. The color scale indicates CNV states: blue shades represent potential deletions and red shades denote possible amplifications.
